# Adaptive immunity to SARS-CoV-2 in cancer patients: The CAPTURE study

**DOI:** 10.1101/2020.12.21.20248608

**Authors:** Annika Fendler, Lewis Au, Laura Amanda Boos, Fiona Byrne, Scott T.C. Shepherd, Ben Shum, Camille L. Gerard, Barry Ward, Wenyi Xie, Maddalena Cerrone, Georgina H. Cornish, Martin Pule, Leila Mekkaoui, Kevin W. Ng, Richard Stone, Camila Gomes, Helen R. Flynn, Ana Agua-Doce, Phillip Hobson, Simon Caidan, Mike Howell, Robert Goldstone, Mike Gavrielides, Emma Nye, Bram Snijders, James Macrae, Jerome Nicod, Adrian Hayday, Firza Gronthoud, Christina Messiou, David Cunningham, Ian Chau, Naureen Starling, Nicholas Turner, Jennifer Rusby, Liam Welsh, Nicholas van As, Robin Jones, Joanne Droney, Susana Banerjee, Kate Tatham, Shaman Jhanji, Mary O’Brien, Olivia Curtis, Kevin Harrington, Shreerang Bhide, Tim Slattery, Yasir Khan, Zayd Tippu, Isla Leslie, Spyridon Gennatas, Alicia Okines, Alison Reid, Kate Young, Andrew Furness, Lisa Pickering, Sonia Ghandi, Steve Gamblin, Charles Swanton, on behalf of the Crick COVID19 consortium, Emma Nicholson, Sacheen Kumar, Nadia Yousaf, Katalin Wilkinson, Anthony Swerdlow, Ruth Harvey, George Kassiotis, Robert Wilkinson, James Larkin, Samra Turajlic, on behalf of the CAPTURE consortium

**Affiliations:** Cancer Dynamics Laboratory, The Francis Crick Institute, London, NW1 1AT, UK; Skin and Renal Units, The Royal Marsden NHS Foundation Trust, London, SW3 6JJ, UK; Tuberculosis Laboratory, The Francis Crick Institute, London, NW1 1AT, UK; Retroviral Immunology Laboratory, The Francis Crick Institute, London, NW1 1AT, UK; Research Department of Haematology at University College London Cancer Institute, WC1E 6DD, London, UK; Autolus Limited, The MediaWorks, 191 Wood Lane, London, W12 7F; Experimental Histopathology Laboratory, The Francis Crick Institute, London, NW1 1AT, UK; Mass Spectrometry Proteomics Science Technology Platform, The Francis Crick Institute, London, NW1 1AT, UK; Flow Cytometry Scientific Technology Platform, The Francis Crick Institute, London, NW1 1AT, UK; Safety, Health & Sustainability, The Francis Crick Institute, London, NW1 1AT, UK; Advances Sequencing Facility, The Francis Crick Institute, London, NW1 1AT, UK; Scientific Computing Scientific Technology Platform, The Francis Crick Institute, London, NW1 1AT, UK; Metabolomics Scientific Technology Platform, The Francis Crick Institute, London, NW1 1AT, UK; Immunosurveillance Laboratory, The Francis Crick Institute, London, NW1 1AT, UK; Department of Pathology, The Royal Marsden NHS Foundation Trust, London, NW1 1AT, UK; Department of Radiology, The Royal Marsden NHS Foundation Trust, London, SW3 6JJ, UK; Gastrointestinal Unit, The Royal Marsden NHS Foundation Trust, London, SW3 6JJ, UK; Breast Unit, The Royal Marsden NHS Foundation Trust, London, SW3 6JJ, UK; Neuro-oncology Unit, The Royal Marsden NHS Foundation Trust, London, SW3 6JJ, UK; Clinical Oncology Unit, The Royal Marsden NHS Foundation Trust, London, SW3 6JJ, UK; Sarcoma Unit, The Royal Marsden NHS Foundation Trust, London, SW3 6JJ, UK; Palliative Medicine, The Royal Marsden NHS Foundation Trust, London, SW3 6JJ, UK; Gynaecology Unit, The Royal Marsden NHS Foundation Trust, London, SW3 6JJ, UK; Anaesthetics, Perioperative Medicine and Pain Department, The Royal Marsden NHS Foundation Trust, London, SW3 6JJ, UK; Lung Unit, The Royal Marsden NHS Foundation Trust, London, SW3 6JJ, UK; Head and Neck, The Royal Marsden NHS Foundation Trust, London, SW3 6JJ, UK; Acute Oncology Service, The Royal Marsden NHS Foundation Trust, London, SW3 6JJ, UK; Neurodegeneration Biology Laboratory, The Francis Crick Institute, London, NW1 1AT, UK; Structural Biology of Disease Processes Laboratory, The Francis Crick Institute, London, NW1 1AT, UK; Cancer Evolution and Genome Instability Laboratory, The Francis Crick Institute, London, NW1 1AT, UK; Haemato-oncology Unit, The Royal Marsden NHS Foundation Trust, London, SW3 6JJ, UK; Division of Genetics and Epidemiology and Division of Breast Cancer Research, The Institute of Cancer Research, London, SW7 3RP, UK; Worldwide Influenza Centre, The Francis Crick Institute, London, NW1 1AT, UK; Department of Infectious Disease, Imperial College London, W12 0NN, UK; The Wellcome Center for Infectious Disease Research in Africa, University Cape Town, Cape Town, Observatory 7925, Republic of South Africa

**Keywords:** SARS-CoV-2, COVID-19, Cancer, Adaptive Immunity, Antibody Response, Neutralising Antibodies, T-cell Response, Prospective Study, Vaccine

## Abstract

There is a pressing need to characterise the nature, extent and duration of immune response to SARS-CoV-2 in cancer patients and inform risk-reduction strategies and preserve cancer outcomes. CAPTURE is a prospective, longitudinal cohort study of cancer patients and healthcare workers (HCWs) integrating longitudinal immune profiling and clinical annotation. We evaluated 529 blood samples and 1051 oronasopharyngeal swabs from 144 cancer patients and 73 HCWs and correlated with >200 clinical variables. In patients with solid cancers and HCWs, S1-reactive and neutralising antibodies to SARS-CoV-2 were detectable five months post-infection. SARS-CoV-2-specific T-cell responses were detected, and CD4^+^ T-cell responses correlated with S1 antibody levels. Patients with haematological malignancies had impaired but partially compensated immune responses. Overall, cancer stage, disease status, and therapies did not correlate with immune responses. These findings have implications for understanding individual risks and potential effectiveness of SARS-CoV-2 vaccination in the cancer population.

## INTRODUCTION

Severe acute respiratory syndrome coronavirus 2 (SARS-CoV-2) associated with coronavirus disease 2019 (COVID-19) has resulted in a global pandemic with a particularly detrimental impact on cancer patients. Concern that cancer patients may be more susceptible to SARS-CoV-2 infection and severe COVID-19 led to broad risk-mitigation strategies such as strict self-shielding, reduced cancer disease monitoring, and modification of cancer treatment (Neal, Nekhlyudov, Wheatstone and Koczwara, 2020; Richards, Anderson, Carter, Ebert and Mossialos, 2020; Schrag, Hershman and Basch, 2020). Such strategies, which largely lack evidence, might have a profoundly negative impact on cancer outcomes. Critically, little weight was given to the heterogeneity of cancer as a disease, or the varying impact of cancer treatments on host immunity. To date, higher COVID-19 mortality was reported in patients with lung and haematological cancers compared with other tumour types (Garassino et al., 2020; García-Suárez et al., 2020; Kuderer et al., 2020; Lee et al., 2020), and in those with progressive cancer versus those in remission, irrespective of cancer type (Kuderer et al., 2020). The precise impact of systemic anti-cancer therapy (SACT), radiotherapy and cancer surgery on the course and outcome of SARS-CoV-2 infection is debated (2020; Liu et al., 2020; Luo, Rizvi, Egger, Preeshagul, Wolchok and Hellmann, 2020; Robilotti et al., 2020). As routine SARS-CoV-2 screening has not been widely adopted, the true prevalence and changing incidence of SARS-CoV-2 infection amongst cancer patients remains unknown, and outcomes of those with asymptomatic or mild infection are underrepresented in studies to date (Saini, Tagliamento, Lambertini, McNally, Romano, Leone, Curigliano and de Azambuja, 2020). Advanced age, comorbidities (i.e. diabetes and hypertension), smoking, and obesity, which are linked to severe COVID-19, are common in cancer patients (Albiges et al., 2020; Williamson et al., 2020), further highlighting the complexity of interactions in this population. Finally, both diagnosis and management of suspected/confirmed COVID-19 are complicated by the overlap of symptoms with those of the underlying malignancy and/or treatment side effects.

Calibration of current and future risk-mitigation measures, including vaccine effectiveness, will also require an understanding of the impact of cancer and cancer treatments on the nature, extent and duration of immunity to SARS-CoV-2. In non-cancer patients, SARS-CoV-2 infection results in an antibody response to major antigens (spike and nucleocapsid proteins) as well as an neutralising antibody response (Moderbacher et al., 2020; Ripperger et al., 2020; Wajnberg et al., 2020a). The magnitude of the response appears to be dependent on disease severity and persists up to 6 months, based on reports so far (Moderbacher et al., 2020; Robbiani et al., 2020; Wajnberg et al., 2020a). As well as having a direct antiviral effect (Mathew et al., 2020), CD4^+^ T-cells are critical for inducing antibody response and B-cell memory to SARS-CoV-2 infection (Ni et al., 2020). Emerging data indicate that SARS-CoV-2-specific memory T-cells are maintained beyond six months following infection (Sekine et al., 2020). A retrospective study in cancer patients reported lower seroconversion rates in 10 cancer patients compared with 14 healthcare workers (HCWs) (30% vs. 71%) at 15 days post onset of COVID-19 disease (POD) (Solodky, Galvez, Russias, Detourbet, N’Guyen-Bonin, Herr, Zrounba and Blay). The cohort size is limited but raised the possibility of delayed or failed seroconversion in some cancer patients. Prospective evaluation and data on neutralising antibody or T-cell responses are lacking in cancer patients. The exact correlates of longer-term immune protection from SARS-CoV-2 remain unknown in the general population, especially as reports of reinfection are limited (Tillett et al.; To et al., 2020; Van Elslande et al., 2020). Nevertheless, both antibody and T-cell responses are induced by SARS-CoV-2 vaccines (Jackson et al., 2020a; Karlsson, Humbert and Buggert, 2020; Krammer, 2020; Lederer et al.; Walsh et al., 2020), the effectiveness of which remains unexplored in cancer patients. This, coupled with the uncertain impact of vaccination on transmission, cancer patients are again faced with the prospect of continued self-shielding and modified treatments under sweeping advice for individuals who are “extremely vulnerable”, even after SARS-CoV-2 vaccination (JCVI, 2020).

CAPTURE (COVID-19 antiviral response in a pan-tumour immune monitoring study; NCT03226886) is a prospective, longitudinal cohort study initiated in response to the global SARS-CoV-2 pandemic and its impact on cancer patients (Au, Boos, Swerdlow, Byrne, Shepherd, Fendler and Turajlic, 2020). The study recruits cancer patients with and without SARS-CoV-2 infection to evaluate the impact of cancer and cancer therapies on the immune response to SARS-CoV-2. Given cancer patients’ exceptional reliance on the healthcare system and high risk of nosocomial infection, a related aim of the study is to evaluate viral prevalence, shedding, clearance, and anti-viral immunity in HCWs. This is relevant to transmission dynamics in the hospital setting. Here we present a planned interim analysis relating to cohort characteristics, antibody, and T-cell responses to SARS-CoV-2 in 144 cancer patients and 73 HCWs.

## RESULTS

### CAPTURE study design and cohort characteristics

Inclusion criteria for CAPTURE study were intentionally broad, and patients were recruited irrespective of cancer type, stage, or treatment. Patients in Group A had suspected or confirmed, current or past SARS-CoV-2 infection (**Figure 1; STAR Methods**). Patients in Group B were recruited in the course of routine clinical care (**Figure 1**), without suspected or confirmed SARS-CoV-2 infection, past or present. Critically, inclusion of this group allows us to identify undiagnosed *asymptomatic* infections. At each study visit we collected combined oronasopharyngeal swabs, plasma, serum, and peripheral blood mononuclear cells (PBMC), as well as data relating to COVID-19, cancer, comorbidities, and medication (**STAR Methods**). Patients completed digital questionnaires (PROFILES https://profiles-study.rmh.nhs.uk/; **STAR Methods**) on lifestyle, self-shielding practice, occupational and household exposure to SARS-CoV-2, and quality-of-life. The same study procedures were applied in HCWs and data were self-reported via a questionnaire (**Figure 1; STAR Methods**).

**Figure 1:**
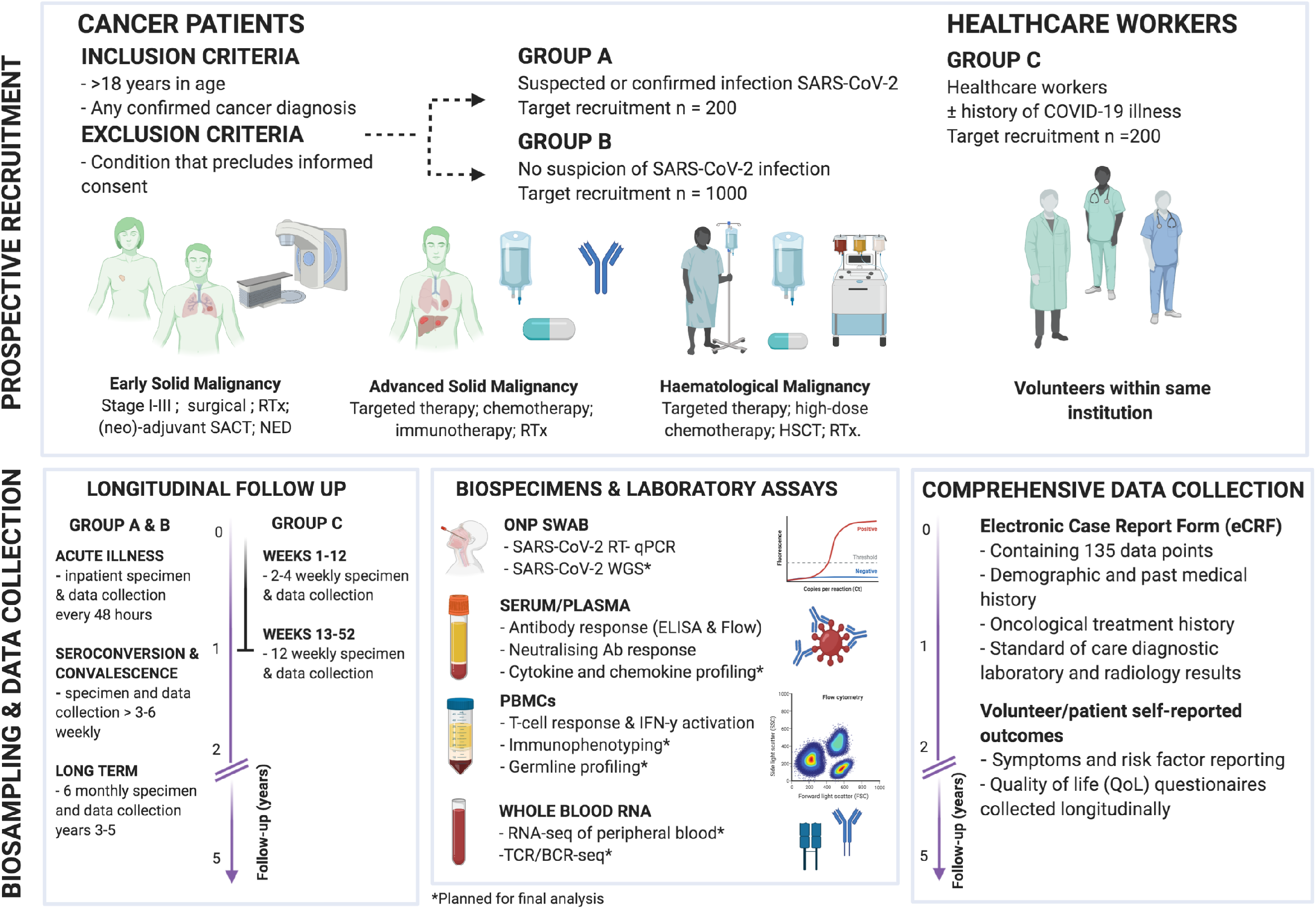
CAPTURE recruitment, follow up schedules, sample, and data collection overview, and planned analyses. Cancer patients irrespective of cancer type, stage, or treatment were recruited into two study arms: Group A: patients with suspected or confirmed, current or past SARS-CoV-2 infection; Group B: patients without suspected or confirmed SARS-CoV-2 infection. Group C: healthcare workers (HCWs) within the same institution. Follow up schedules for cancer patients were bespoke to their COVID-19 status and account for their clinical schedules (inpatients: every 2 – 14 days; outpatients: every clinical visit maximum every 3-6 weeks in year one and every six months in year two, and at the start of every or every-second cycle of treatment (**STAR Methods**). HCWs were enrolled under a one-year follow up schedule as shown. Clinical data, oronasopharyngeal swabs and blood were collected at each study visit. Viral antigen testing (RT-PCR on swabs), antibody (ELISA, flow cytometric assay), T-cell response and IFN-γ activation assays were performed. Additional planned analyses for the final analysis are shown. *BCR - B-cell receptor; ONP - Oronasopharyngeal; ELISA - enzyme-linked immunoassay; PBMC - peripheral blood mononuclear cells; TCR - T-cell receptor; WGS - whole genome sequencing*

144 cancer patients were enrolled in the CAPTURE study from May to October 2020 (74 in Group A, and 70 in Group B). The median follow-up time was 135 days (range: 8-202 days). The median age was 59 (range 28-82), and 48% of the patients were male (**Table S1**). One or more comorbid conditions were present in 67 (46%) patients, including: diabetes (10%), hypertension (26%), obesity (body mass index (BMI) ≥30; 17%), and autoimmune/inflammatory disease (8%) (**Table S2**). 31% of the patients were current smokers and 49% ex-smokers. 134 (93%) patients had a diagnosis of solid malignancy; the majority had stage IV disease (**Table S2**). Ten (7%) patients had a diagnosis of a haematological malignancy, of which half were in remission (**Table S2**). 113 (78%) patients received SACT in the 12 weeks prior to enrolment: 46 (25%) with immune checkpoint inhibitors (CPIs) and 57 (31%) with chemotherapy. 18 (10%) patients received radiation therapy, and eight (4%) patients had surgery in the 12 weeks prior to study enrolment. With respect to the most recent anti-cancer intervention for patients with advanced disease, 31% of patients had disease progression, 61% were responding to treatment or had stable disease, and 7% had no evidence of active disease. Cancer treatment had been delayed (deferral of at least one SACT cycle or radiotherapy fraction or surgery) in 31% and modified in 6% as a consequence of the SARS-CoV-2 pandemic (**Table S2**).

73 HCWs were enrolled in the same period. Median follow-up time was 82 days (range: 1-97) (**Table S1**). Compared with cancer patients, HCWs were generally younger (94% were <65 years), more likely to be female (77%), and had fewer comorbidities. 27% of HCWs were ex-smokers and only one HCW was a current smoker. The majority (91%) worked in patient-facing roles (**Table S3**).

### SARS-CoV-2 case definition in cancer patients

In order to achieve an accurate case definition with respect to current or prior SARS-CoV-2 infection we performed SARS-CoV-2 RT-PCR and antibody testing in all patients, irrespective of clinical presentation (**Figure 2; STAR Methods**). Patients were swabbed at each study visit and results of any clinically indicated SARS-CoV-2 RT-PCR tests were also noted. 74 cancer patients had presented with suspected COVID-19 (Group A), of which 35/74 (47%) had a positive SARS-CoV-2 RT-PCR (**Table 1**). In patients with serial positive swabs (n=15), median duration of viral shedding was 11 days (range: 4-60 days), while three patients had intermittently positive swabs in the course of active COVID-19. Rather than re-infection, this likely reflects false negative RT-PCR results as reported in the literature (Woloshin, Patel and Kesselheim, 2020). 70 patients were recruited without suspicion of current or prior SARS-CoV-2 infection (Group B), and none had a positive SARS-CoV-2 RT-PCR result to date.

**Table 1:** Baseline characteristics and clinical course in laboratory confirmed SARS-CoV-2 patients and in patients with clinical suspicion of COVID-19 but negative for SARS-CoV-2.

|  | Confirmed<br>SARS-CoV2 positive<sup>a</sup> | Group A,<br>SARS-CoV2 negative<sup>a</sup> |  |
| --- | --- | --- | --- |
| Clinical Characteristics | n= 43<sup>b</sup> | n= 33 | p-value<sup>c</sup> |
| Age, years (median, range) | 59.4 (38- 78) | 57.6 (28- 78) | 0.539 |
| Male | 21 (49) | 14 (42) | 0.746 |
| **Smoking status** |  |  |  |
| Current smoker | 15 (37) | 12 (36) |  |
| Ex-smoker | 18 (44) | 12 (36) | 0.664 |
| Never smoker | 5 (12) | 7 (21) |  |
| Unknown | 3 (7) | 2 (6) |  |
| **Past medical history<sup>d</sup>** |  |  |  |
| HTN | 10 (24) | 12 (36) | 0.411 |
| PVD/IHD/CVD | 3 (7) | 12 (36) | 0.254 |
| Diabetes Mellitus | 8 (19) | 7 (21) | 0.207 |
| Obese | 11 (15) | 8 (11) | 1 |
| Inflammatory/Autoimmune | 1 (2) | 2 (6) | 0.758 |
| **Cancer stage<sup>e</sup>** |  |  |  |
| Stage I-II | 2 (5) | 2 (6) |  |
| Stage III | 7 (16) | 3 (9) | 0.792 |
| Stage IV | 27 (63) | 24 (73) |  |
| Haematological | 5 (12) | 4 (12) |  |
| **COVID-19 illness** |  |  |  |
| **Symptoms<sup>d</sup>** |  |  |  |
| Fever | 34 (79) | 22 (67) | 0.339 |
| Dyspnoea | 24 (59) | 13 (39) | 0.234 |
| Cough | 28 (65) | 10 (30) | **0.005** |
| Fatigue | 8 (17) | 9 (27) | 0.368 |
| Anosmia | 8 (19) | 2 (6) | 0.207 |
| Myalgia | 13 (30) | 6 (18) | 0.349 |
| Coryzal illness | 6 (14) | 6 (18) | 0.854 |
| Headache | 9 (21) | 6 (18) | 0.993 |
| Diarrhoea | 11 (26) | 4 (12) | 0.241 |
| Asymptomatic | 2 (5) | 1 (3) | 0.719 |
| Other reason for presentation, yes | 6 (20) | 26 (79) | **<0.001** |
| Duration of symptoms, median (range) | 19 (1- 146) | - | - |
| **Admission to hospital** |  |  |  |
| Not hospitalised | 19 (44) | - | - |
| Admitted with COVID-19-like illness | 20 (47) | - | - |
| Developed COVID-19 during admission | 4 (9) | - | - |
| Duration of admission, median (range) | 8 (1- 120) | - | - |
| Required supplemental oxygen, yes | 14 (33) | - | - |
| WHO Score<sup>f</sup>, median (range) | 3 (1- 7) | - | - |
| Admission to ICU, yes | 4 (9) | - | - |
| **Complications of COVID-19** |  |  |  |
| Pneumonia | 7 (16) | - | - |
| Required corticosteroids (>10 mg equivalent prednisolone) | 5 (10) | - | - |
| Need for mechanical ventilation/NIV | 3 (7) | - | - |
| Venous thromboembolism | 1 (2) | - | - |
| **Clinical diagnostics** |  |  |  |
| **Radiological abnormalities** |  |  |  |
| **CXR<sup>h</sup>** |  |  |  |
| normal | 9 (21) | - | - |
| abnormal in keeping with COVID | 12 (28) | - | - |
| abnormal, indeterminate | 3 (7) | - | - |
| abnormal | 3 (7) | - | - |
| NA | 16 (37) | - | - |
| **CT<sup>h</sup>** |  |  |  |
| normal | 2 (4.7) | - | - |
| abnormal in keeping with COVID | 8 (19) | - | - |
| abnormal, indeterminate | 2 (5) | - | - |
| abnormal | 2 (5) | - | - |
| NA | 29 (67) | - | - |
| **Laboratory Investigations** |  |  |  |
| **FBC** |  |  |  |
| Hb, < 120 g/L | 0 (0) | - | - |
| WBC, >11 x 10<sup>9</sup>/L | 1 (3) | - | - |
| NO, <2.0 x 10<sup>9</sup>/L | 11 (29) | - | - |
| NO, >7.5 x 10<sup>9</sup>/L | 2 (5) | - | - |
| Plt, <150 x 10<sup>9</sup>/L | 11 (28) | - | - |
| **Biochemistry** |  |  |  |
| Creatinine, >1.5 ULN | 0 (0) | - | - |
| CRP, > 10g/dl | 26 (74) | - | - |
| **Microbiology** |  |  |  |
| SARS-CoV2 PCR positive | 35 (81) | - | - |
| Duration of PCR positivity, median (range) | 11 (4- 60) | - | - |
| **Other respiratory pathogen detected** |  |  |  |
| Respiratory/viral screen native | 37 (86) | - | - |
| Respiratory viral pathogen | 1 (2) | - | - |
| Respiratory bacterial pathogen | 1 (2) | - | - |
| NA screen | 4 (9) | - | - |

CRP, C-reactive protein; CT, computed tomography scan; CVD, cerebrovascular disease; CXR; chest x-ray; Hb, haemoglobin; HTN, hypertension; ICU, intensive care unit; IHD, ischaemic heart disease; IQR, interquartile range; LLN, lower limit of normal; NIV, non-invasive ventilation; NO, neutrophils; PCR, polymerase chain reaction; Plt, platelets; PVD, peripheral vascular disease; ULN, upper limit of normal; WBC, white blood cell.
a, Indicates positive/negative for SARS-CoV2 by laboratory definition; b, Expressed as number and percent of affected individuals unless otherwise specified. Denominators reflect available data; unless specified, unknowns are not included; c, Significance tests: Chi-squared test of categorical variables and Mann-Whitney U test for comparison of median values; d, Consensus from patient reported outcomes and/or review of electronic patient records; e, Cancer stage at enrolment according to AJCC stage definition (**STAR Methods**) ; f, WHO COVID-19 severity score (**STAR Methods**); g, The primary cause of death in this patient was advanced cancer and not related to complications of COVID-19; h, Scored according to British Thoracic Imaging Society COVID guidance (**STAR Methods**).

**Figure 2:**
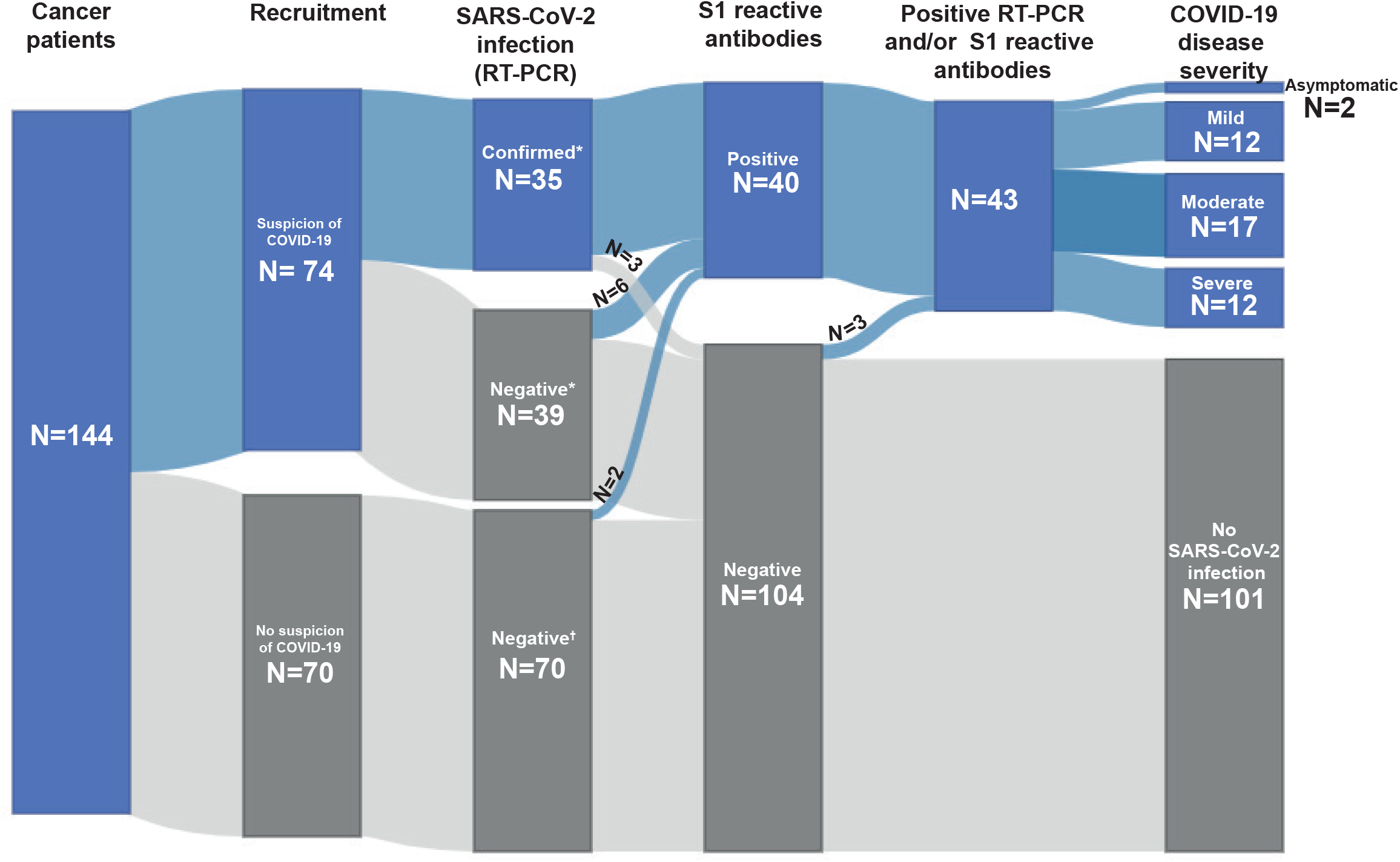
SARS-CoV-2 infection and antibody status in cancer patients. Distribution of SARS-CoV-2 infection, and S1-reactive antibody status and COVID-19 severity in cancer patients. 144 cancer patients were recruited between May and October 2020 with either a suspicion of COVID-19 or no suspicion of COVID-19. SARS-CoV-2 infection status by RT-PCR and S1-reactive antibodies were analysed at recruitment and in serial samples. RT-PCR results prior to recruitment were extracted from electronic patient records. COVID-19 case definition includes all patients with either RT-PCR confirmed SARS-CoV-2 infection or S1-reactive antibodies. *RT-PCR to confirm SARS-CoV-2 infection was performed at symptom onset for all cases with suspicion of COVID-19 and serially at each follow-up visit; three cases had a positive RT-PCR result at the time of recruitment, 32 cases had a confirmed SARS-CoV-2 infection prior to recruitment, 39 cases tested negative despite suspicion of COVID-19. ^+^RT-PCR to confirm SARS-CoV-2 infection was performed at study recruitment and serially at each follow-up visit.

Next, we quantified total S1-reactive antibody titres by ELISA in 144 patients at multiple time points, with at least one sample taken at >25 days after POD (**Figure 2; STAR Methods**). A total of 326 serum samples were tested, median of 2 (range 1-10) per patient. We detected S1-reactive antibodies in 32/35 (91%) patients with RT-PCR confirmed SARS-CoV-2 infection; 6/39 (15%) patients with suspected COVID-19 accompanied by typical radiological changes, but negative SARS-CoV-2 RT-PCR; and 2/70 patients (3%) with no suspicion of COVID-19 and negative SARS-CoV-2 RT-PCR at study entry. Therefore, 43/144 (30%) of cancer patients had confirmed SARS-CoV-2 infection by RT-PCR or S1-reactive antibody, or both (**Figure 2**).

94 (65%) patients completed the digital questionnaire (**STAR Methods**), allowing us to evaluate potential sources of SARS-CoV-2 exposure. 82/94 (85%) had been advised to self-shield and all reported adherence to the guidance (**Table S4**). Six patients with confirmed SARS-CoV-2 infection, and zero in the SARS-CoV-2 negative group reported that a member of their household had confirmed COVID-19 (*P*<0.001) (**Table S4**).

### Clinical correlates of COVID-19 diagnosis and severity in cancer patients

In the 43 patients with confirmed SARS-CoV-2 infection, fever (n=34, 79%), cough (n=28, 65%), dyspnoea (n=24, 59%), myalgia (n=13, 30%), and diarrhoea (n=11, 26%) were the most common presenting symptoms (**Figure S1A**). The median number of symptoms was 3 (range 0-7). 20 (47%) patients were asymptomatic by day 28 POD, while eight (19%) patients remained symptomatic >50 days POD with a maximum of 231 days POD. In 33/74 patients (45%) COVID-19 was suspected but excluded by SARS-CoV-2 RT-PCR and antibody. We note that these patients were less likely to present with cough, but just as likely to present with fever or dyspnoea as those with confirmed SARS-CoV-2 infection, (*P*<0.01; **Table 1**), highlighting the challenge of clinical case definition of SARS-CoV-2 in cancer patients. Alternative diagnoses were found in 26/33 (79%) patients: non-respiratory infection (biliary sepsis, urinary tract infection, gastroenteritis, postoperative infection or neutropenic sepsis) in 11 (33%); progressive disease and cancer related symptoms in six (18%); respiratory infection without a confirmed pathogen in three (9%); mycoplasma pneumoniae in two (6%); CPI-induced pneumonitis in two (6%); other pathologies (gastroesophageal reflux, cerebrovascular event) in two (6%); adverse effects of medication in one (3%).

Of the patients with confirmed COVID-19 who were symptomatic, 12/41 (30%) had mild, 17 (41%) moderate, and 12 (29%) severe illness, according to the WHO severity score system (**STAR Methods**). 20 (47%) patients were hospitalised due to COVID-19, 14 (33%) required supplemental oxygen, four (9%) were admitted to an intensive care unit (ICU), with one requiring mechanical ventilation and inotropic support (**Tables 1 and S5**). Five (12%) patients were treated with corticosteroids (>10mg prednisolone equivalent), and two (5%) patients with haematological malignancy and severe COVID-19 received tocilizumab, an anti-IL-6 monoclonal antibody. One patient had a thrombo-embolic complication. The median duration of in-patient stay was eight days (range 1-120 days), and all patients recovered. At database lock (October 2020), four (10%) patients with confirmed SARS-CoV-2 infection died of progressive cancer, and no patients died due to established complications of COVID-19 (**Table 1**). 26 (61%) patients with SARS-CoV-2 infection had had subsequent cancer treatment delayed and four (9%) had restarted with dose or therapy modification (**Table S2**).

Compared with patients with solid malignancies, patients with haematological malignancies were at higher risk of ICU admission (OR: 3.8, 95% CI 2.65-1090; **STAR Methods**) (**Table S6**). Otherwise, we found no significant association between risk of severe disease or hospital admission with cancer type, stage, or disease status after recent SACT, nor sex, age, obesity, smoking status or comorbidities across all cancer patients (**Table S6**). The size of the SARS-CoV-2 positive cohort limits subgroup analyses in this interim report.

### SARS-CoV-2 antibody response in cancer patients

Forty patients, 38 with symptomatic COVID-19 and two without symptoms, had SARS-CoV-2 S1-reactive antibodies (**Figure 3A**). Three patients who were positive by SARS-CoV-2 RT-PCR had no evidence of S1-reactive antibodies on serial testing 25-150 days after POD. They include a patient with diffuse large B cell lymphoma (DLBCL) and severe COVID-19 who had received anti-CD19 CAR-T cells three months prior to presentation; a patient with Hodgkin’s lymphoma and mild COVID-19 illness who had received ABVD chemotherapy one month prior to presentation; and a patient with metastatic colorectal carcinoma (mCRC) and mild COVID-19 who was on FOLFIRI chemotherapy at the time of presentation. All other patients including those with a diagnosis of leukaemia had evidence of S1-reactive antibodies (**Figure 3B**).

**Figure 3:**
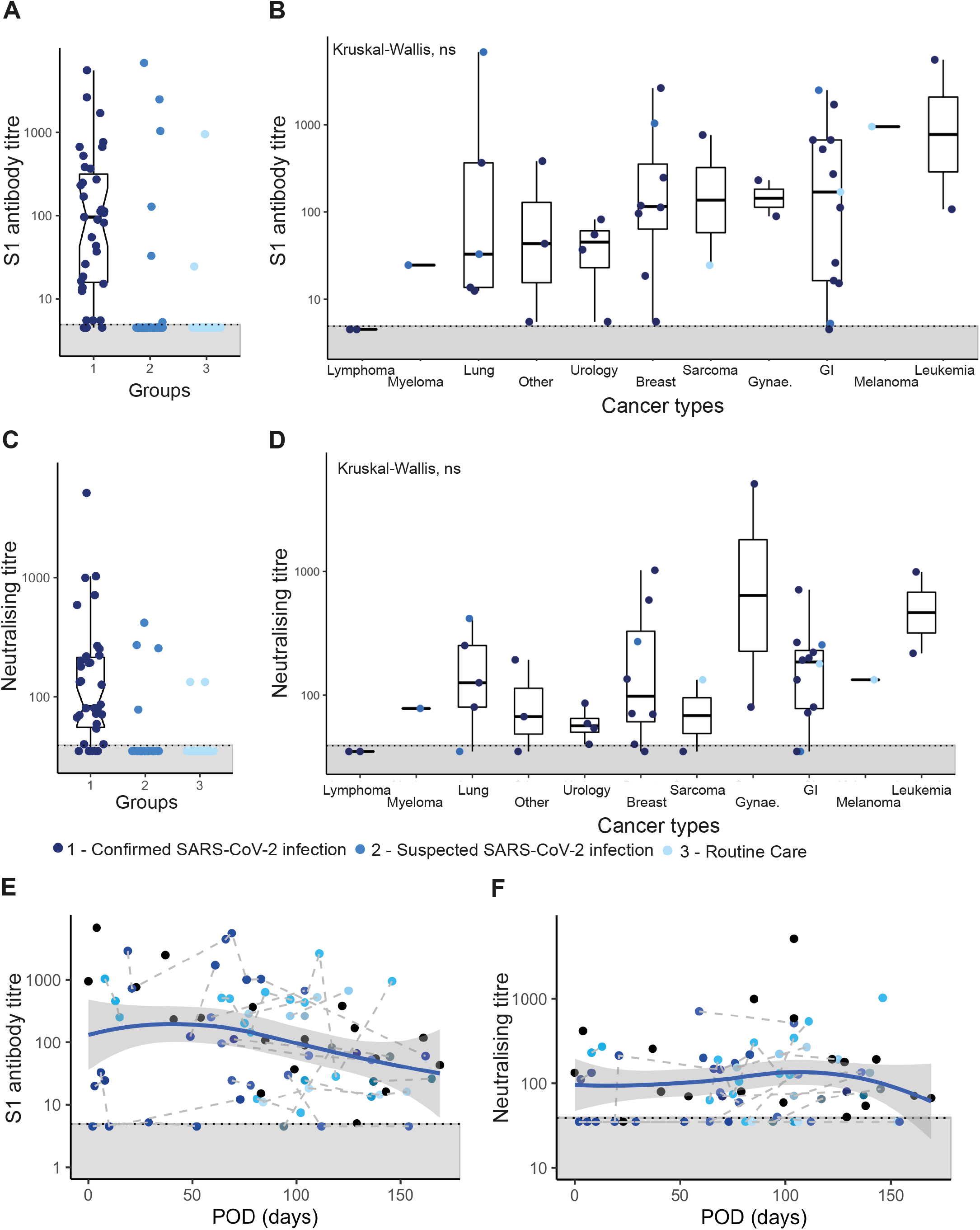
Antibody response in cancer patients. (A) S1-reactive antibodies were measured in serial serum dilutions with titres representing reciprocal serum dilutions. Cancer patients were grouped into confirmed SARS-CoV-2 infection (n=35), suspected SARS-CoV-2 infection (n=39) and recruited during routine care (n=70). (B) S1-reactive antibody titres across cancer types. (C) Neutralising titres were measured by live virus neutralisation assays in Vero-E6 cells and are displayed as reciprocal serum dilutions. (D) Neutralising titres across cancer types. Dots indicate individual patient values. Boxes show median as well as upper and lower quantiles. Notches (if present) indicate the 95%CI of the median. Whiskers represent the 1.5 times IQR. B) S1-reactive antibody titres across cancer types. (E) S1-reactive antibody titres and (F) neutralising titres over time after POD. Black dots denote patients with only one serial sample, coloured dots denote patients with serial samples. Loess regression was performed and is displayed as a blue line with 95% CI as grey area. Dotted lines and grey areas at graph bottom indicate limit of detection.

We conducted sensitive flow cytometric assays to quantify spike-reactive (S) IgG, IgM and IgA levels in sera of all patients with S1-reactive antibodies (**Figure S2A and S3A, STAR Methods**). S-specific IgG was detected in 38 patients (**Figure S2B**) and IgM in 23 patients (**Figure S2C**), with levels significantly correlated with S1-reactive antibody titres (*P*<0.0001) (**Figure S2E-F**). The lower proportion of those with IgM response are in keeping with reports in non-cancer patients in the convalescent phase (Röltgen et al., 2020; Zohar et al.). S-reactive IgA was detected in serum of only four (10%) patients (**Figure S2D**), reflecting either decline of IgA after the acute phase of infection (Rodda et al., 2020), or competitive IgG binding in the flow assay (Ng et al., 2020).

We next evaluated whether patients’ sera could neutralise SARS-CoV-2 in Vero E6 cultures. We observed neutralising activity in 34/42 patients (**Figure 3C-D**). Of the eight patients with no evidence of neutralising activity, three had no S1-reactive antibodies and three had S1-reactive antibodies in the lower quantile (titre ≤25). Neutralising titres were significantly correlated with S1-reactive antibody titres (*P* < 0.0001)(**Figures S2G**).

Beside the significant findings in patients with haematological malignancies we did not observe any correlation between S1-reactive and neutralising antibody response with cancer characteristics (**Figure S4A**). S1-reactive antibody titres correlated with COVID-19 severity (**Figure S4B**) as previously reported in non-cancer patients (Chen et al., 2020; Wang et al., 2020). S1-reactive antibody titres also significantly correlated with age (**Figure S4C**) and presence of diabetes mellitus, neither of which correlated with COVID-19 disease severity in this study. We did not observe a significant correlation of disease severity with neutralising antibody titres, in contrast to studies in non-cancer patients (Chen et al., 2020; Wang et al., 2020). Presence of neutralising antibodies was not associated with cancer characteristics and patient demographics (**Table S6**).

Peak antibody response to SARS-CoV-2 infection is usually detected ∼30 days after POD, followed by a decline (Long et al., 2020; Seow et al., 2020) and then plateau of antibody titres (Ripperger et al., 2020; Wajnberg et al., 2020a). We evaluated serial serum samples in 20 cancer patients with a median of two time points per patient (range 1-10), and median follow-up time of 127 days after POD (range: 15-176). 19/20 had S1-reactive antibodies at the time of enrolment (median 84 days after POD, range: 4-171) (**Figure 3E**). One patient had no antibodies at enrolment (94 days after POD) but seroconverted at day 128 after POD. Neutralising antibodies were detected as early as day eight (**Figure 3F**), and as late as day 171 after POD, in agreement with reports in non-cancer patients (Dan et al., 2020). S1-reactive antibody titres showed a weak declining trend, while neutralising titres remained stable up to 176 days. S1-reactive and neutralising titres declined below detection limit in only one patient with metastatic breast cancer and mild COVID-19, at 118 days after POD.

Given that cross-reactivity to seasonal human coronaviruses (HCoVs) (Ng et al., 2020) has been reported we evaluated matched pre-pandemic sera from 47 patients, 10 with and 37 without S1-reactive antibodies. We found no evidence of S1-reactive antibodies in the pre-pandemic sera in any patient (**Figure S2H**), but S-reactive IgG or IgM were detected in 18 patients without S1-reactive antibodies (**Figure S2B-C**). These observations are consistent with cross-reactivity induced by prior HCoV infection against S, but not S1 antigen (Ng et al., 2020).

### SARS-CoV-2 case definition in HCWs

Of 73 HCWs, nine had prior RT-PCR confirmed SARS-CoV-2 infection, eight in the context of symptomatic presentation, and one via staff-screening program (**Figure 4A; STAR Methods**). All HCWs had serial SARS-CoV-2 RT-PCR tests per the CAPTURE protocol, and none tested positive in the course of follow up (median 82 days from enrolment, range 1-97). We quantified total S1-reactive antibody titres by ELISA in 203 plasma samples (median: 3 per patient, range: 1-4) collected within 20 to 185 days after POD. S1-reactive antibodies were detected in 8/9 HCWs with prior RT-PCR confirmed SARS-CoV-2; 6/24 HCWs with a past history of symptoms consistent with COVID-19, but no contemporaneous RT-PCR test; and 7/40 HCWs without any history of COVID-19 symptoms and negative RT-PCR at study entry (**Figure 4B**). Therefore, 22/73 HCWs (30%) had confirmed SARS-CoV-2 infection by RT-PCR or S1-reactive antibody or both. Fourteen had been symptomatic and 8 asymptomatic, consistent with previous reports of frequent asymptomatic infections in HCWs (Houlihan et al., 2020). There was no significant association between the presence of symptoms and age, sex, smoking status, BMI, comorbidities, or place of work (**Table S7**). We observed a non-significant trend towards association between S1-reactive antibody titres and the presence of symptoms (*R*^2^=0.39, *P*=0.09) (**Figure S4E**).

**Figure 4:**
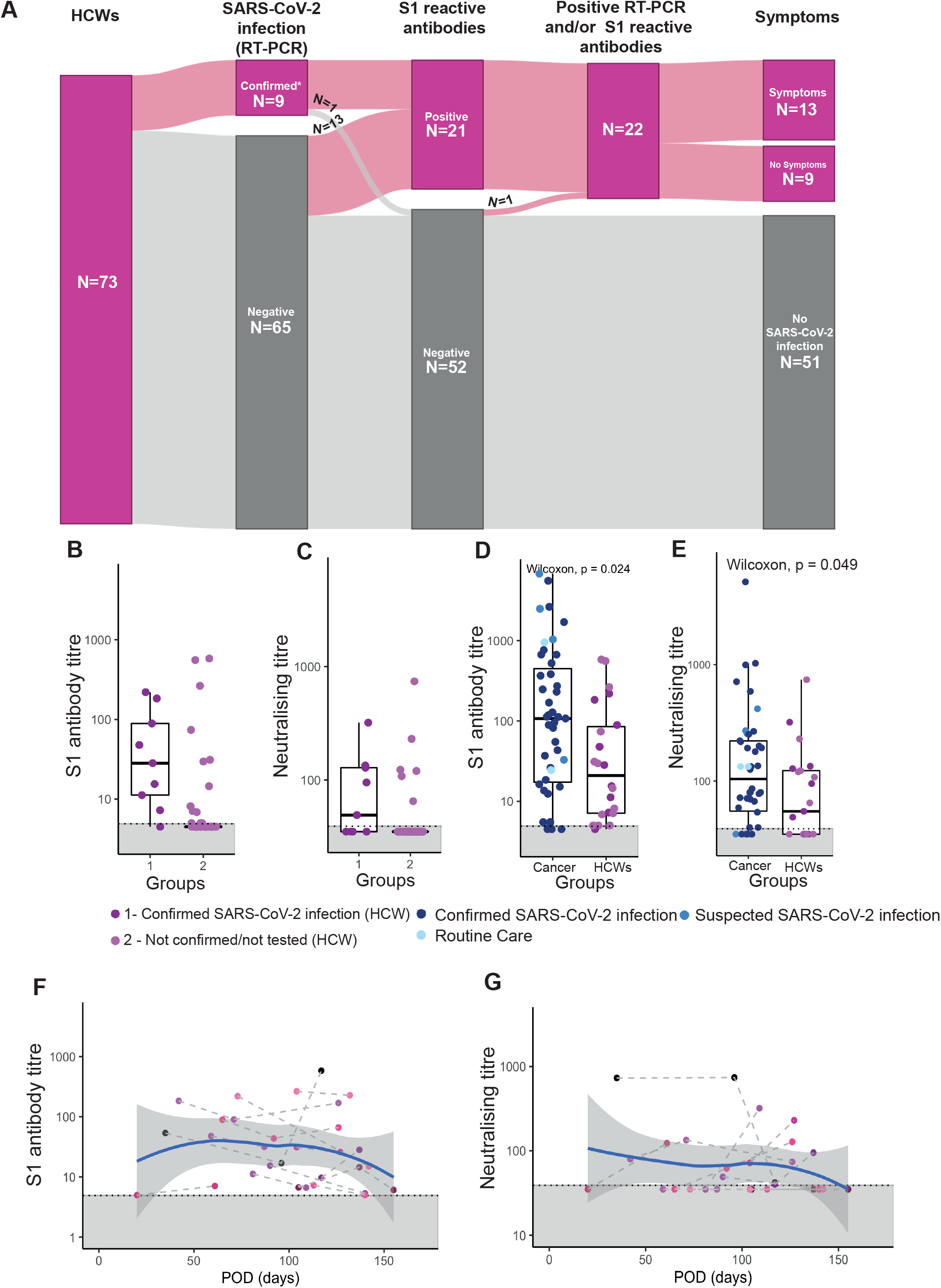
Antibody response in HCWs. (A) Distribution of SARS-CoV-2 infection, and S1-reactive antibody status and reported symptoms in HCWs. 73 HCWs were recruited between May and October 2020, all with unknown SARS-CoV-2 infection status. SARS-CoV-2 infection status by RT-PCR and S1-reactive reactive antibodies were analysed at recruitment and in serial samples. RT-PCR results prior to recruitment were extracted from participant questionnaires. COVID-19 case definition includes all patients with either RT-PCR confirmed SARS-CoV-2 infection or S1-reactive antibodies. (B) S1-reactive antibodies (C) and neutralising antibodies were measured in serial serum dilutions with titres representing reciprocal serum dilutions. HCWs were grouped into confirmed SARS-CoV-2 infection (n=9), and negative or no previous PCR test (n=64). (D) S1-reactive and (E) neutralising antibody titres in cancer patients and HCWs. Dots represent individual patients. Boxes represent upper and lower quantiles; the line represents the median. Whiskers represent 1.5 x IQR. Colors denote recruitment groups of cancer patients and HCWs. (F) S1-reactive antibody titres and (G) neutralising titres over time after POD. Black dots denote HCWs with only one serial sample, coloured dots denote HCWs with serial samples. Loess regression was performed and is displayed as a blue line with 95% CI as grey area. Dotted lines and grey areas at graph bottom indicate limit of detection.

With respect to potential occupational exposure, only working in ICU appeared to confer a higher risk of SARS-CoV-2 infection in this cohort of HCWs (**Table S3**). There was no difference in household exposure between HCWs with and without SARS-CoV-2 infection (**Table S4**).

Headache (n=12, 60%), anosmia (n=10, 50%), myalgia (n=10, 50%), fever (n=10, 50%), and cough (n=9, 45%) were the most commonly reported symptoms at presentation in HCWs (**Figure S1B**). Anosmia, headache and coryzal illness were more common presentations in HCWs compared to cancer patients, while fever was more frequent in cancer patients (*P*<0.05, **Table S8**). Most HCWs (n=15, 75%) were asymptomatic at 14 days after POD, but three remain symptomatic with cough and dyspnoea >50 days after POD. No HCWs were hospitalised due to COVID-19. Duration of symptoms did not differ significantly between HCWs and cancer patients (**Table S8**).

S-specific IgG and IgM were detected in 18/21 and 10/21 HCWs with S1-reactive antibodies, respectively (**Figure S5A-B**). IgG levels were significantly correlated with S1-reactive antibody titres (**Figure S5D-E**). IgA was detected in one (4.8%) HCW (**Figure S5C**). S-specific IgG or IgM were also detected in five HCWs without S1-reactive antibodies, again suggesting cross-reactivity against S, but not S1 after prior HCoV infection.

Neutralising antibodies were detected in 58% of HCWs with S1-reactive antibodies, compared with 87% of cancer patients (*P*<0.05) (**Figure 4C**). Neutralising titres were significantly correlated with S1-reactive antibody titres (**Figure S5F**). Presence of neutralising antibodies did not differ significantly between HCWs with or without symptoms (p = 0.6). Overall, both S1-reactive and neutralising titres were lower in HCWs compared with cancer patients (**Figure 4D-E**). We evaluated serial samples (median two timepoints) in 13 HCWs, where POD was apparent, with a median follow-up time of 127 days after POD (range: 61-155 days). S1-reactive and neutralising antibody titres were below the detection limit in baseline samples in three cases, at 75, 81, and 82 days after POD, respectively but all seroconverted by day 104, 113, and 117, respectively (**Figure 4F-G**). We observed a weak declining trend for both S1-reactive and neutralising antibodies, but both remained detectable at maximum follow up of 155 days.

### SARs-CoV-2-specific T-cells are detected in cancer patients and HCWs

CD4^+^ and CD8^+^ T-cells contribute to the antiviral immune response through activation of plasma and memory B-cells, and also have a direct antiviral effect (Karlsson, Humbert and Buggert, 2020). We performed in vitro stimulation of PBMC with synthetic SARS-CoV-2 peptide pools, consisting of 15-mer sequences with 11 amino acid overlap covering the immunodominant parts of the spike (S) protein and the complete sequence of the nucleocapsid (N) and membrane (M) proteins in 27/43 patients and 20/21 HCWs with confirmed SARS-CoV-2 infection (**Figure 5A and S3B, STAR Methods**). SARs-CoV-2-specific CD4^+^ T-cells and CD8^+^ T-cells (SsT-cells) (identified by activation induced markers OX40, CD137, and CD69 (Grifoni et al.)) were measured at a single time-point, a median of 85 days after POD (range: 2-128 days). We detected CD4^+^ T-cells in 20/27 (74%), and CD8^+^ T-cells and 16/27 (59%) of patients (**Figure 5B-C**). Consistent with observations in non-cancer patients (Houlihan et al., 2020; Moderbacher et al., 2020; Weiskopf et al., 2020), percentage of CD4^+^ T-cells after stimulation consistently predominated over CD8^+^ T-cells (**Figure S6A**). The highest levels of SARS-CoV-2-specific CD4^+^ and CD8^+^ T-cells were observed in the patients with lymphoma who lacked an S1-reactive antibody response. We detected no SsT-cells in two patients with leukaemia where both had evidence of S1-reactive antibody. One patient with T-cell acute lymphoblastic leukaemia had received an allogeneic stem cell transplant and alemtuzumab (anti-lymphocyte monoclonal antibody) 59 days after POD. The other, with the diagnosis of acute myeloid leukaemia, was receiving anthracycline and cytarabine chemotherapy at the time of diagnosis of SARS-CoV-2 infection. SARS-CoV-2-specific T-cells were not detected in four patients with solid malignancies (mNSCLC, mBC, mGOJ, mCRC), who had evidence of S1-reactive antibodies, but two lacked neutralising antibodies.

**Figure 5:**
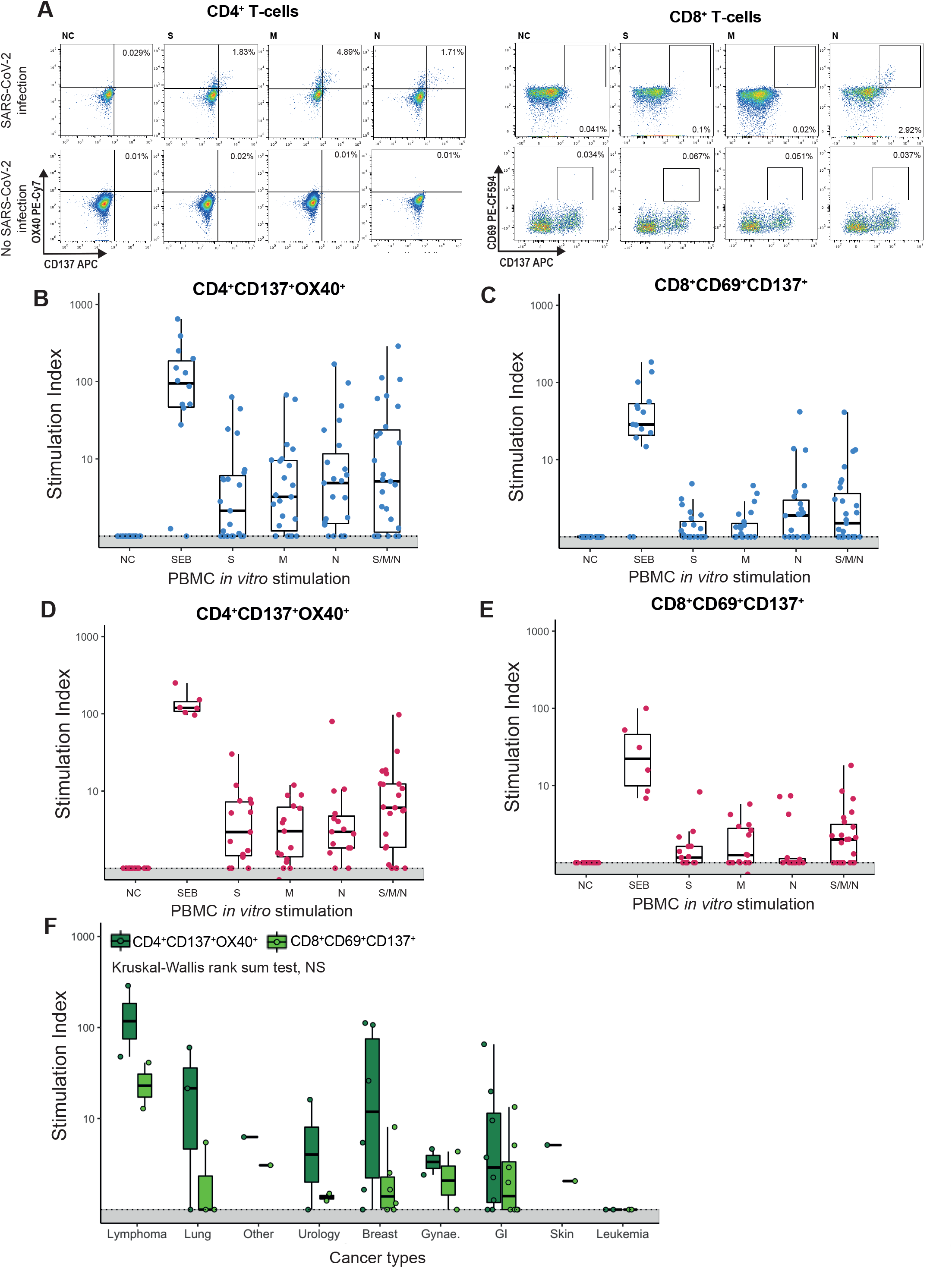
T-cell response in cancer patients and HCWs. (A) Representative plots of CD4^+^ and CD8^+^ T-cells in a patient with confirmed COVID-19 and a cancer patient without COVID-19 after in vitro stimulation with S, M, and N peptide pools, positive control (Staphylococcal enterotoxin B, SEB) or negative control (NC). Frequency of Sars-CoV-2-specific (B) CD4^+^ and (C) CD8^+^ T-cells in cancer patients and (D) CD4^+^ and (E) CD8^+^ T-cells in HCWs. Stimulation index was calculated by dividing the percentage of positive cells in the stimulated sample by the percentage of positive cells in the negative control (NC). To obtain the total number of SsT-cells the sum of cells activated by S, M, and N was calculated (SMN). (F) CD4^+^ and CD8^+^ cells T-cells across cancer types. Individual patient’s data are represented as dots. Boxes mark the upper and lower quantiles, and line indicates the median. Whiskers represent 1.5 x IQR. Dotted lines and grey areas at graph bottom indicate limit of detection.

We detected SARS-CoV-2-specific CD4^+^ and CD8^+^ T-cells in 17/20 and 11/20 HCWs, respectively (**Figure 5D-E**). The percentage of CD8^+^ T-cells after stimulation was lower than percentage of CD4^+^ T-cells (**Figure S6B**), however one HCW had evidence of CD8^+^ T-cells but no SARS-CoV-2 reactive CD4^+^ T-cells (although neutralising antibodies were present). We noted a non-significant trend to lower levels of SsT-cells in cancer patients who received anti-PD-1 therapy in the three months prior to POD (**Figure S4D**), but no other significant correlations (**Figure S4A**).

IFN-⍰, secreted by SsT-cells (Crotty, 2019), was detected after *in vitro* stimulation with levels correlated to the number of SsT-cells (**Figure S6C**). There was no difference between the proportion of HCWs and cancer patients with SsT-cells (78% vs. 85%, *P*>0.05), nor the percentage of SsT-cells (**Figure S6D-E**).

Both in the acute and convalescent phase of SARS-CoV-2 infection, a significant proportion of SARS-CoV-2-specific CD4^+^ T-cells are T follicular helper cells (Tfh) (Juno et al., 2020; Moderbacher et al., 2020), which are required for IgG and neutralising response by B-cells (Murugesan et al., 2020). In our cohort, following stimulation with S-specific peptide pools the number of CD4^+^ T-cells was significantly correlated with S1-reactive antibody titres, suggesting the latter may reflect Tfh T-cell activation (**Figure S6F-H**).

Cross-reactive T-cell responses to HCoVs are observed frequently in healthy individuals (Grifoni et al.; Mateus et al., 2020). We extended the T-cell assay to 12 cancer patients and HCWs without confirmed SARS-CoV-2 infection. Cross-reactive CD4^+^ T-cells were detected in 7/12 and CD8^+^ T-cells in 3/12 participants, but the overall T-cell percentage was significantly lower than those in subjects with confirmed infections (*P*<0.05) (**Figure S6I-J**).

## DISCUSSION

This is the first report from a prospective study of the adaptive immune response to SARS-CoV-2 in cancer patients and HCWs. Recruitment to CAPTURE commenced in May 2020, which marked the end of the first wave of SARS-CoV-2 infections in the UK. As a result, most participants in this report were infected prior to study enrolment and evaluated in the convalescent phase. Thanks to broad inclusion criteria, enhanced laboratory criteria for case definition, and longitudinal follow up, we provide an accurate picture of SARS-CoV-2 status and dynamics of the anti-viral immune response, in the context of clinical presentation of COVID-19 and cancer characteristics. In this interim analysis our data indicate that most cancer patients with symptomatic COVID-19 mount a functional adaptive immune response to SARS-CoV-2 infection, which appears durable for at least five months. HCWs, who on the whole experienced milder illness and were not well matched to the cancer cohort, had lower S1-reactive and neutralising antibody levels, but similar antibody kinetics and a comparable SARS-CoV-2 T-cell response.

The small number of patients with haematological malignancies is an important exception characterised by an impaired immune response to SARS-CoV-2 infection, which may be the basis of higher mortality reported in this group ^5,7^. Haematological malignancies offer a unique opportunity to dissect the contribution of specific immune lineages to infection response (Vardhana and Wolchok, 2020). In our cohort, two patients with B-cell lymphoma had no evidence of antibody response, two patients with leukaemia lacked SsT-cells, and one patient with myeloma had low titres of S1-reactive and neutralising antibodies. These findings are attributable to a number of interrelated factors - lineage defects driven by the underlying malignancy, therapy (i.e., CAR-T therapy, allogeneic stem cell transplant, myeloablative chemotherapy), and therapy complications (i.e., GVHD). While it appears in these examples that defects in antibody or T-cell response are compensated by the other adaptive immunity arm, patients with haematological malignancies were still at higher risk of ICU admission (albeit based on small numbers), and two received tocilizumab, which may have aided their recovery. All the patients in our cohort recovered, while 11/18 patients with haematological malignancies died due to COVID-19 at our institution (Angelis et al., 2020), before enrolment into CAPTURE commenced. Thus, it is possible that the patients with haematological malignancy in our interim analysis cohort are not entirely representative of the spectrum of immune dysfunction in this group, which we continue to interrogate through ongoing recruitment and posthumous access to stored samples from those who died.

Most patients with solid malignancies mounted S1-reactive and neutralising antibody responses that appeared stable and comparable to what has been reported in non-cancer populations. There were no unifying features in the small number of patients with solid cancers who lacked antibody or T-cell response. We observed delayed seroconversion in one cancer patient and three HCWs, in line with reports that patients with mild to moderate COVID-19 experience slower seroconversion rates (Bar-Or et al., 2020; Wajnberg et al., 2020a; Wajnberg et al., 2020b). While precise correlates of immune-protection to SARS-CoV-2 remain elusive, SARS-CoV-2 vaccine efforts are largely focused on induction of neutralising antibodies, non-human primate studies show that they are protective (Mercado et al., 2020) and effective vaccines all induce a robust neutralising response (Anderson et al., 2020; Folegatti et al., 2020; Jackson et al., 2020b; Polack et al., 2020; Voysey et al.; Walsh et al., 2020). Cancer patients are not represented in current SARS-CoV-2 vaccine studies but will nevertheless be prioritised in the vaccination programme (JCVI, 2020). Although our data suggests cancer patients on the whole are no less likely to seroconvert in response to SARS-CoV-2 infection, attenuated effectiveness and seroconversion of some vaccines such as influenza have been reported in cancer setting (i.e. anti-CD20 blockade) (Bar-Or et al., 2020; Beck, McKenzie, Hashim, Harris and Nguyen-Van-Tam, 2012). Further, mRNA-based vaccines (Jackson et al., 2020b; Sahin et al., 2020), which are delivered as liposomal nanoparticles, may in theory suffer from passive sequestration of liposomes into tumour tissue (Fanciullino, Ciccolini and Milano, 2020), something that has been exploited in anti-cancer therapy (i.e. liposomal doxorubicin) (Green and Rose, 2006; Sahin et al., 2020). Overall, it remains an aim of our program to evaluate vaccine response in CAPTURE participants.

We also detected IFN-y expressing SsT-cells in the majority of cancer patients and HCWs, with the number of S-specific CD4^+^ T-cells correlated with the S1-reactive antibody response, consistent with their role in B-cell activation. In the context of the outbreak of SARS in 2003, SARS-specific T-cells were detected up to 17 years after infection, much longer than antibodies (Le Bert et al., 2020). The finding of lower proportion of patients with SARS-CoV-2-specific CD8^+^ T-cells, compared to those with CD4^+^ T-cells, and their overall lower numbers, may reflect the bias of our peptide pool design with 15-mer peptide pools, optimised for MHC Class II presentation and preferentially recognised by CD4^+^ T-cells (Grifoni et al.). In addition, CD8^+^ T-cell responses are induced by a broader range of antigens than CD4^+^ T-cells, specifically epitopes within non-structural proteins that are underrepresented in our pool covering the immunodominant regions of S, N, and M (Grifoni et al.).

There are limitations in this report, many of which will be addressed with longer follow up and planned analyses within the CAPTURE program. Our data so far reflect those who recovered from COVID-19 but included a broad representation of illness from asymptomatic to severe. We have in the majority recruited convalescent patients, and hence this interim report is focused on the immune protective response. Ongoing work will explore correlates of immunopathology during acute infection.

These data provide insights into the branches of adaptive immunity against SARS-CoV-2, placed in the context of cancer and cancer therapy, which also bear relevance for the effectiveness of SARS-CoV-2 vaccines. Further questions that can only be assessed with longer follow up are the impact of treatment interruptions during the first wave of the pandemic on long term cancer outcomes, and the impact on quality of life for those with persistent symptoms following SARS-CoV-2 infection (Yelin et al., 2020). The prospective, longitudinal framework of this study integrating clinical outcomes and immune profiling will serve to generate data rapidly to address these and other emergent questions as the pandemic evolves, including immune responses to new SARS-CoV-2 variants (Kemp et al., 2020), and during the ensuing vaccination programme.

## Data Availability

Request for further information or for resources and reagents should be directed and will be fulfilled by the lead contact, Samra Turajlic

## ACKNOWLEDGMENTS

We thank the CAPTURE trial team, including Eleanor Carlyle, Kim Edmonds, and Lyra Del Rosario, as well as Somya Agarwal, Hamid Ahmod, Natalie Ash, Ravinder Dhaliwal, Lauren Dowdie, Tara Foley, Lucy Holt, Justine Korteweg, Charlotte Lewis, Karla Lingard, Mary Mangwende, Aida Murra, Kema Peat, Sarah Sarker, Nahid Shaikh, Sarah Vaughan, and Fiona Williams. We acknowledge the tremendous support from the clinical and research teams at participating units at the Royal Marsden Hospital, including Ethel Black, Arnold Dela Rosa, Carole Pearce, Jessica Bazin, Leonora Conneely, Chloe Burrows, Tommy Brown, Jeremy Tai, Emma Lidington, Holly Hogan, Amanda Upadhyay, David Capdeferro, Ingrid Potyka, Annette Drescher, Farzana Baksh, Melissa Balcorta, Catia Da Costa Mendes, Joao Amorim, Venus Orejudos, and Louise Davison. We also thank the Volunteer Staff at The Francis Crick Institute, the Crick COVID19 consortium, Alice Lilley for help with neutralising assays, Antonia Toncheva, and the cloning unit at Autolus including James Sillibourne, Katarzyna Ward, Katarina Lamb and Philip Wu. We thank Brigitta Stockinger for her thoughtful review and comments on the manuscript. A.F. has received funding from the European Union’s Horizon 2020 research and innovation programme under the Marie Skłodowska-Curie grant agreement No. 892360. L.A. is funded by the Royal Marsden Cancer Charity. Due to the pace at which the field is evolving, we acknowledge researchers in COVID-19, particularly in furthering our understanding of SARS-CoV-2 infection and we apologise for work that was not cited. This research was funded in part, by the Cancer Research UK (grant reference number C50947/A18176). The Francis Crick Institute receives its core funding from Cancer Research UK (CRUK) (FC010110), the UK Medical Research Council (FC010110), and the Wellcome Trust (FC010110). For the purpose of Open Access, the author has applied a CC BY public copyright licence to any Author Accepted Manuscript version arising from this submission. TRACERx Renal is partly funded by the National Institute for Health Research (NIHR) Biomedical Research Centre (BRC) at the Royal Marsden Hospital and Institute of Cancer Research (ICR) (A109). The CAPTURE study is sponsored by The Royal Marsden NHS Foundation Trust and funded from a grant from The Royal Marsden Cancer Charity.

## AUTHOR CONTRIBUTIONS

Conceptualisation, S.T., L.A. and L.A.B.; Methodology, S.T., A.Fendler, F.B., K.W., G.K. and R.H. Software, M.G.; Formal Analysis, A.Fendler., L.A., S.T.S., G.K., K.W., R.H. and M.C.; Investigation, A.F., L.A., F.B., S.T.C.S, B.S., C.G., W.X., B.W., K.W., M.C., A.A-D. and R.H.; Resources, S.T., A.Fendler, L.A., L.A.B., F.B., S.T.S., B.S., C.G., B.W., W.X., M.C., G.C., M.P. and L.M., R.S., C.G., H.F., M.G., F.G., O.C., T.S., Y.K., Z.T. and I.L.; Data Curation, L.A.B., S.T.S, B.S., C.G., A.Fendler and L.A.; Writing – Original Draft: S.T., A.Fendler, L.A., S.T.S.; Writing - Review & Editing: All; Visualization, A.Fendler, S.T.C.S, S.T. and L.A.; Supervision, S.T.; Trial conduct, S.T., L.A., L.A.B., J.L., N.Y., A.R., E.N. and S.K.

## DECLARATION OF INTERESTS

S.T. is funded by Cancer Research UK (grant reference number C50947/A18176), the National Institute for Health Research (NIHR) Biomedical Research Centre at the Royal Marsden Hospital and Institute of Cancer Research (grant reference number A109), the Kidney and Melanoma Cancer Fund of The Royal Marsden Cancer Charity, The Rosetrees Trust (grant reference number A2204), Ventana Medical Systems Inc (grant reference numbers 10467 and 10530), the National Institute of Health (US) and the Melanoma Research Alliance. S.T. has received speaking fees from Roche, Astra Zeneca, Novartis and Ipsen. S.T. has the following patents filed: Indel mutations as a therapeutic target and predictive biomarker PCTGB2018/051892 and PCTGB2018/051893 and Clear Cell Renal Cell Carcinoma Biomarkers P113326GB. J.L. has received research funding from Bristol-Myers Squibb, Merck, Novartis, Pfizer, Achilles Therapeutics, Roche, Nektar Therapeutics, Covance, Immunocore, Pharmacyclics, and Aveo, and served as a consultant to Achilles, AstraZeneca, Boston Biomedical, Bristol-Myers Squibb, Eisai, EUSA Pharma, GlaxoSmithKline, Ipsen, Imugene, Incyte, iOnctura, Kymab, Merck Serono, Nektar, Novartis, Pierre Fabre, Pfizer, Roche Genentech, Secarna, and Vitaccess. C.S. acknowledges grant support from Pfizer, AstraZeneca, Bristol Myers Squibb, Roche-Ventana, Boehringer-Ingelheim, Archer Dx Inc (collaboration in minimal residual disease sequencing technologies) and Ono Pharmaceutical, is an AstraZeneca Advisory Board member and Chief Investigator for the MeRmaiD1 clinical trial, has consulted for Pfizer, Novartis, GlaxoSmithKline, MSD, Bristol Myers Squibb, Celgene, AstraZeneca, Illumina, Genentech, Roche-Ventana, GRAIL, Medicxi, Bicycle Therapeutics, and the Sarah Cannon Research Institute, has stock options in Apogen Biotechnologies, Epic Bioscience, GRAIL, and has stock options and is co-founder of Achilles Therapeutics. Patents: C.S. holds European patents relating to assay technology to detect tumour recurrence (PCT/GB2017/053289); to targeting neoantigens (PCT/EP2016/059401), identifying patent response to immune checkpoint blockade (PCT/EP2016/071471), determining HLA LOH (PCT/GB2018/052004), predicting survival rates of patients with cancer (PCT/GB2020/050221), identifying patients who respond to cancer treatment (PCT/GB2018/051912), a US patent relating to detecting tumour mutations (PCT/US2017/28013) and both a European and US patent related to identifying insertion/deletion mutation targets (PCT/GB2018/051892). C.S. is Royal Society Napier Research Professor (RP150154). His work is supported by the Francis Crick Institute, which receives its core funding from Cancer Research UK (FC001169), the UK Medical Research Council (FC001169), and the Wellcome Trust (FC001169). C.S. is funded by Cancer Research UK (TRACERx, PEACE and CRUK Cancer Immunotherapy Catalyst Network), Cancer Research UK Lung Cancer Centre of Excellence, the Rosetrees Trust, Butterfield and Stoneygate Trusts, NovoNordisk Foundation (ID16584), Royal Society Research Professorship Enhancement Award (RP/EA/180007), the NIHR BRC at University College London Hospitals, the CRUK-UCL Centre, Experimental Cancer Medicine Centre and the Breast Cancer Research Foundation, USA (BCRF). His research is supported by a Stand Up To Cancer-LUNGevity-American Lung Association Lung Cancer Interception Dream Team Translational Research Grant (SU2C-AACR-DT23-17). Stand Up To Cancer is a program of the Entertainment Industry Foundation. Research grants are administered by the American Association for Cancer Research, the Scientific Partner of SU2C. C.S. also receives funding from the European Research Council (ERC) under the European Union’s Seventh Framework Programme (FP7/2007-2013) Consolidator Grant (FP7-THESEUS-617844), European Commission ITN (FP7-PloidyNet 607722), an ERC Advanced Grant (PROTEUS) from the European Research Council under the European Union’s Horizon 2020 research and innovation programme (835297) and Chromavision from the European Union’s Horizon 2020 research and innovation programme (665233). L.P. has received research funding from Pierre Fabre, and honoria from Pfizer, Ipsen, Bristol-Myers Squibb, and EUSA Pharma. A.F., L.A., L.A.B., F.B., S.T.C.S., A.S., B.S, BW, CG, WX have no conflicts of interest to declare.

## STAR METHODS

### RESOURCE AVAILABILITY

#### Materials Availability

Materials used in this study will be made available upon request. There are restrictions to the availability based on limited quantities.

#### Data and software availability

Original data will be made available upon request.

## EXPERIMENTAL MODEL AND SUBJECT DETAILS

### Study design

CAPTURE is a prospective, longitudinal study, the study design has previously been described in detail (Au, Boos, Swerdlow, Byrne, Shepherd, Fendler and Turajlic, 2020). The primary and secondary endpoints of the study relate to the impact of COVID-19 on long term survival outcomes. Exploratory endpoints pertain to characterising clinical and immunological determinants of COVID-19 in cancer patients. Planned interim analysis took place after 6 months of active recruitment. CAPTURE was approved as a substudy of TRACERx Renal (NCT03226886). TRACERx Renal was initially approved by NRES Committee London Fulham on 17/01/2012. The TRACERx Renal sub-study CAPTURE was submitted as part of Substantial Amendment 9 and approved by London - Fulham Research Ethics Committee on 01/05/2020 and the Health Research Authority on 30/04/2020. CAPTURE is performed in accordance with the ethical principles in the Declaration of Helsinki, Good Clinical Practice and applicable regulatory requirements.

### Study Population, Study schedule and follow-up

Eligible cases in the cancer patient cohort include adult patients with current or history of invasive cancer who were recruited either into group A (with suspected or confirmed SARS-CoV-2 infection) or group B (no clinical indication to test for SARS-CoV-2 or negative RT-PCR for SARS-CoV-2). Healthcare workers without a history of cancer are eligible for group C. Study exclusion criterion is medical or psychological conditions that precludes consent. Study recruitment commenced on 04/05/2020 and is ongoing.

Clinical data and sample collection for participating cancer patients is performed at baseline, and at clinical visits per standard-of-care management during the first year of follow up, frequency varies depending on in- or outpatient status and systemic anti-cancer treatment regimens. For inpatients, study assessments are repeated every 2 – 14 days. For outpatients, the follow-up study assessments are aligned with clinically indicated hospital attendances. The frequency of study assessments in the first year for patients on anti-cancer therapies are as follows: every cycle for patients on immune checkpoint inhibitors or targeted therapies; every second cycle for patients receiving chemotherapy; every outpatient appointment (maximum 6 weekly) for patients on endocrine therapy or in surveillance or routine cancer care follow up. Patient reported data is collected 3-monthly via an online questionnaire. In year two to five of follow up, the frequency of study assessments is reduced (see Supplementary Material Study Protocol). For healthcare workers, study sampling and self-reported data are collected via an online questionnaire at baseline and repeated 2-4 weekly for three months and 3-monthly thereafter for a follow up period of one year.

### Data and Sample Sources

Patient-reported outcome data are collected using PROFILES (Patient Reported Outcomes Following Initial treatment and Long-term evaluation of Survivorship; https://profiles-study.rmh.nhs.uk/). PROFILES is a web-based questionnaire administration and management system designed for the study of the physical and psychosocial impact of cancer and its treatment. Online questionnaires for baseline and follow up assessments were designed to record data for cancer patients and healthcare workers participating in CAPTURE. Self-reported data collected for cancer patients includes ethnicity, smoking status, alcohol consumption, recent travel history, occupation, exercise habits, dietary habits, previous medical history, autoimmune disease (self, next of kin), vaccination history, concomitant medication, self-shielding status, previous SARS-CoV-2 tests, SARS-CoV-2 tests in household members, current and recent symptoms. Additional data collected for healthcare workers include gender, age, patient facing professional role, hospital area of placement, placement in COVID-19 dedicated area, exposure to aerosols, taking SARS-CoV-2 swabs from patients. For cancer patients, data on quality-of-life metrics using a European Organisation for Research and Treatment of Cancer (EORTC) QLQ-C30 questionnaire as per study schedule. Further demographic, epidemiological and clinical data (e.g., cancer type, cancer stage, treatment history) are collected from the internal electronic patient record system and entered into detailed case report forms in a secure electronic database. Findings on chest imaging (chest X-ray and CT thorax) are collected and classified based on the guidance for reporting radiologists by the British Society of Thoracic Imaging (BSTI) (BSTI, 2020) into “abnormal – in keeping with COVID-19” (comprising BSTI categories “classic COVID-19” and “probable COVID-19”), indeterminate for COVID-19 (corresponding to BSTI category “indeterminate for COVID-19”), “abnormal – non-COVID-19” and “normal” (corresponding to BSTI category “non-COVID”).

Study samples collected comprise blood samples, oropharyngeal swabs and archival and excess material from routine clinical investigations. Detailed sampling schedule and methodology has been previously described (Au, Boos, Swerdlow, Byrne, Shepherd, Fendler and Turajlic, 2020). Surplus serum from patient biochemistry samples taken as part of routine care were also retrieved and linked to the study IDs before anonymisation and study analysis. Collected data and study samples are de-identified and stored with only the study-specific study identification number. For self-reported data, a PROFILES member number, which is generated automatically.

### Cell lines and viruses

SUP-T1 cells stably transfected with spike or control vectors were obtained from Martin Pule, and Leila Mekkaoui. Vero E6 cells were from the National Institute for Biological Standards and Control, UK. The SARS-CoV-2 isolate hCoV-19/England/02/2020 was obtained from the Respiratory Virus Unit, Public Health England, UK, and propagated in Vero E6 cells.

## METHOD DETAILS

### Handling of oronasopharyngeal swabs and RNA isolation

SARS-CoV-2 RT-PCR was performed from oronasopharyngeal swabs within the Crick Covid19 testing pipeline (Aitken et al., 2020). The complete standard operating procedure is available on the Covid19 consortium website: https://www.crick.ac.uk/research/covid-19/covid19-consortium. Oronasopharyngeal swabs were collected in VTM medium, frozen within 24 hrs after collection, and stored at -80°C until processing. Oronasopharyngeal swabs were handled in a CL3 laboratory inside a biosafety cabinet using appropriate personal protective equipment and safety measures, which were in accordance with a risk assessment and standard operating procedure approved by the safety, health and sustainability committee at the Francis Crick Institute. In brief, 100 µl of swab vial content was inactivated in 5 M Guanidinium thiocyanate. RNA isolation was completely automated on a Biomek FX using a kit-free, silica bead-based method.

### WHO classification of severity of COVID-19

We classified severity of COVID-19 according to the WHO (World Health Organisation) clinical progression scale (Aitken et al., 2020). Uninfected: uninfected, no viral RNA detected - 0; Mild (ambulatory): asymptomatic, viral RNA detected - 1 / symptomatic, independent - 2 / symptomatic, assistance needed - 3; Moderate (hospitalised): no oxygen therapy (If hospitalised for isolation only, record status as for ambulatory patient) - 4 / oxygen by mask or nasal prongs - 5; Severe (hospitalised): oxygen by non-invasive ventilation or high flow – 6 / intubation and mechanical ventilation, pO_2_/FiO_2_ ≥ 150 or SpO_2_/FiO_2_ ≥ 200 - 7 / mechanical ventilation, pO_2_/FiO_2_ < 150 (SpO_2_/FiO_2_ < 200) or vasopressors - 8 / mechanical ventilation, pO_2_/FiO_2_ < 150 and vasopressors, dialysis, or extracorporeal membrane oxygenation - 9; Dead - 10.

### SARS-CoV-2 RT-PCR

SARS-CoV-2 RT-PCR was performed using the real-time fluorescent RT-PCR kit for detecting 2019-nCoV (BGI). All samples were measured in duplicates on separate plates and an internal control was run for each sample. Positive, negative, and extraction controls were included on each plate. Runs were regarded as valid when negative control Ct values were > 37 and positive controls Ct values <37. Samples with non-exponential amplification were excluded from analysis. Samples were only considered positive if Ct values in both runs were <37.

### Handling of whole blood samples

All blood samples and isolated products were handled in a CL2 laboratory inside a biosafety cabinet using appropriate personal protective equipment and safety measures, which were in accordance with a risk assessment and standard operating procedure approved by the safety, health and sustainability committee at the Francis Crick Institute. For indicated experiments, serum or plasma samples were heat-inactivated at 56ºC for 30 min prior to use after which they were used in a CL1 laboratory.

### Plasma and PBMC isolation

Whole blood was collected in EDTA tubes (VWR) and stored at 4ºC until processing. All samples were processed within 24 hrs. Time of blood draw, processing, and freezing was recorded for each sample. Prior to processing tubes were brought to room temperature. PBMC and plasma were isolated by density-gradient centrifugation using pre-filled centrifugation tubes (pluriSelect). Up to 30 ml of undiluted blood was added on top of the sponge and centrifuged for 30 min at 1000 x g at room temperature. Plasma was carefully removed then centrifuged for 10 min at 4000 x g to remove debris, aliquoted and stored at - 80ºC. The cell layer was then collected and washed twice in PBS by centrifugation for 10 min at 300 xg at room temperature. PBMC were resuspended in Recovery cell culture freezing medium (Fisher Scientific) containing 10% DMSO, placed overnight in CoolCell freezing containers (Corning) at -80ºC and then stored at -80ºC.

### Serum isolation

Whole blood was collected in serum coagulation tubes (Vacuette CAT tubes, Greiner) for serum isolation and stored at 4ºC until processing. All samples were processed within 24 hrs. Time of blood draw, processing, and freezing was recorded for each sample. Tubes were centrifuged for 10 min at 2000 x g at 4ºC. Serum was separated from the clotted portion, aliquoted and stored at -80ºC.

### S1-reactive IgG ELISA

Ninety-six-well MaxiSorp plates (Thermo Fisher Scientific) were coated overnight at 4°C with purified S1 protein in PBS (3 μg/ml per well in 50 μl) and blocked for 1 hr in blocking buffer (PBS, 5% milk, 0.05% Tween 20, and 0.01% sodium azide). Sera were diluted in blocking buffer (1:50). Fifty microliters of serum were then added to the wells and incubated for 2 hrs at room temperature. After washing four times with PBS-T (PBS, 0.05% Tween 20), plates were incubated with alkaline phosphatase-conjugated goat anti-human IgG (1:1000, Jackson ImmunoResearch) for 1 hr. Plates were developed by adding 50 μl of alkaline phosphatase substrate (Sigma Aldrich) for 15-30 min after six washes with PBS-T. Optical densities were measured at 405 nm on a microplate reader (Tecan). CR3022 (Absolute Antibodies) was used as a positive control. The cut-off for a positive response was defined as the mean negative value multiplied by 0.35 times the mean positive value.

### Flow cytometry for spike-reactive IgG, IgM, and IgA

SUP-T1 cells were harvested, counted and spike-expressing and control SUP-T1 cells were mixed in a 1:1 ratio. The cell mix was transferred into V-bottom 96-well plates at 20,000 cells per well. Cells were incubated with heat-inactivated sera diluted 1:50 in PBS for 30 min, washed with FACS buffer (PBS, 5% BSA, 0.05% sodium azide), and stained with FITC anti-IgG (clone HP6017, Biolegend), APC anti-IgM (clone MHM-88, Biolegend) and PE anti-IgA (clone IS11-8E10, Miltenyi Biotech) for 30 min (all antibodies diluted 1:200 in FACS buffer). Cells were washed with FACS buffer and fixed for 20 min in 1% PFA in FACS buffer. Samples were run on a Bio-Rad Ze5 analyser running Bio-Rad Everest software v2.4 and analysed using FlowJo v10 (Tree Star Inc.) analysis software. Spike-expressing and control SUP-T1 cells were gated and mean fluorescence intensity (MFI) of both populations was measured. MFI in control SUP-T1 cells was subtracted from MFI in spike-expressing SUP-T1 cells, and resulting values were divided by MFI in control SUP-T1 cells to calculate the specific increase in MFI. Values >2 were considered positive.

### Neutralising antibody assay

Confluent monolayers of Vero E6 cells were incubated with SARS-CoV-2 virus and twofold serial dilutions of heat-treated serum or plasma samples starting at 1:40 for 4 hrs at 37°C, 5% CO2, in duplicates. The inoculum was then removed, and cells were overlaid with viral growth medium. Cells were incubated at 37°C, 5% CO_2_. At 24 hrs post-infection, cells were fixed in 4% paraformaldehyde and permeabilized with 0.2% Triton X-100/PBS. Virus plaques were visualized by immunostaining, as described previously for the neutralisation of influenza viruses using a rabbit polyclonal anti-NSP8 antibody used at 1:1000 dilution and anti-rabbit-HRP conjugated antibody at 1:1000 dilution and detected by action of HRP on a tetramethyl benzidine (TMB) based substrate. Virus plaques were quantified and ID50 was calculated.

### T-cell stimulation

PBMC for in vitro stimulation were thawed at 37 °C and resuspended in 10 ml of warm complete medium (RPMI, 5% human AB serum) containing 0.02% benzonase. Viable cells were counted and 1×106 to 2×106 cells were seeded in 200 µl complete medium per well of a 96-well plate. Cells were stimulated with 4 µl/well PepTivator SARS-CoV-2 S, M, or N pools (representing 1µg/ml final concentration per peptide; Miltenyi Biotec, Surrey, UK). Staphylococcal enterotoxin B (Merck, UK) was used as a positive control at 0.5µg/ml final concentration, negative control was PBS containing DMSO at 0.002% final concentration. PBMC were cultured for 24 hrs at 37°C, 5% CO_2_.

### Activation-induced marker assay

PBMC supernatants were collected for cytokine analysis after stimulation for 24 hrs. Cells were washed twice in warm PBMC. Dead cells were stained with 0.5 µl/well Zombie dye V500 for 15 min at RT in the dark, then washed once with PBS containing 2% FCS (FACS buffer). A surface staining mix was prepared per well, containing 2 µl/well of each antibody for surface staining (see key resources table for a full list of antibodies) in 50:50 brilliant stain buffer (BD) and FACS buffer. PBMC were stained with 50 µl surface staining mix per well for 30 min at RT in the dark. Cells were washed once in FACS buffer and fixed in 1% PFA in FACS buffer for 20 min, then washed once and resuspended in 200 µl PBS. All samples were acquired on a Bio-Rad Ze5 flow cytometer running Bio-Rad Everest software v2.4 and analysed using FlowJo v10 (Tree Star Inc.) analysis software. Compensation was performed with 20 µl antibody-stained anti-mouse Ig, k / negative control compensation particle set (BD Biosciences, UK). 1×10^6^ live CD19-/CD14- cells were acquired per sample. Gates were drawn relative to the unstimulated control for each donor. T-cell response is displayed as a stimulation index by dividing the percentage of AIM-positive cells by the percentage of cells in the negative control. If negative control was 0 the minimum value across the cohort was used. When S, M, and N stimulation were combined the sum of AIM-positive cells was divided by the three times the percentage of positive cells in the negative control.

### IFN-y ELISA

IFN-y ELISA was performed using the human IFN-y DuoSet ELISA (R&D Systems) according to the manufacturer’s instructions. Briefly, 96-well plates were coated overnight with capture antibody, washed twice in wash buffer then blocked with reagent diluent for 2 hrs at room temperature. 100 µl of PBMC culture supernatants were added and incubated for 1 hr at room temperature and washed twice in wash buffer. 100 µl detection antibody diluted in reagent diluent was added per well and incubated for 2 hrs at room temperature. Plates were washed twice in wash buffer. 100 µl streptavidin-HRP dilution was added to the plates and incubated for 20 min in the dark at room temperature, plates were washed twice in wash buffer. Reaction was developed using 200 µl substrate solution for 20 min in the dark at room temperature then stopped with 50 µl stop solution. Optical density was measured at 450 nm on a multimode microplate reader (Berthold). Serial dilutions of standard were run on each plate. Concentrations were calculated by linear regression of standard concentrations ranging (0 - 600) pg/ml and normalized to the number of stimulated PBMC. The assay sensitivity was 5pg/ml.

## QUANTIFICATION AND STATISTICAL ANALYSIS

Data and statistical analysis were done in FlowJo 10 and R v3.6.1 in R studio v1.2.1335 unless otherwise stated. Statistical details for each experiment are provided in the figure legends. Gaussian distribution was tested by Kolmogorov-Smirnov test. Mann-Whitney, Wilcoxon, Kruskal-Wallis, Chi^2^, Fisher’s exact test, and Friedman tests were performed for statistical significance. A p-value < 0.05 was considered significant. The ggplot2 package in R was used for data visualization and illustrative figures were created with BioRender.com. Data are usually plotted as single data points and box plots on a logarithmic scale. For boxplots, boxes represent upper and lower quartiles, line represents median, and whiskers IQR times 1.5. Notches represent confidence intervals of the median. For correlation matrix analysis, spearman rank correlation coefficients were calculated between all parameter pairs using the corrplot package in R without clustering. Spearman rank two-tailed p-values were calculated using the corr.test function within the psych package in R. Benjami-Hochberg correction was performed for multivariate testing. Univariate logistic regression analysis was performed using the glm function with the stats package in R.

## SUPPLEMENTARY INFORMATION

**Figure S1: Overview of symptoms in cancer patients and HCWs with Covid-19 (related to Figure 2)**

A) Symptoms at time of SARS-CoV-2 infection were extracted from electronic patient records. Presence of symptoms in an individual patient is indicated by colored circles. Bars at the left side represent the frequency of symptoms across all cancer patients. For a full list of symptoms in all recruited cancer patients refer to Table 1. B) New or worsening symptoms in HCWs with COVID-19 within three months to recruitment. Presence of symptoms in individual HCWs is indicated by colored circles. Bars at the left side represent the frequency of symptoms across all HCWs. For a full list of symptoms in all recruited HCWs refer to Table S3.

**Figure S2: Antibody response in cancer patients (related to Figure 3)**

(A) Representative plots of S-reactive IgG, IgM and IgA in serum of cancer patients. IgG and IgM are denoted on the axis, IgA positivity is denoted by color scale. S-reactive (B) IgG, (C) IgM, or (D) IgA in cancer patients with confirmed SARS-CoV-2 infection (n=35), suspected SARS-CoV-2 infection (n=39) and recruited during routine care (n=70). Correlation of S1-reactive antibody titres with (E) S-reactive IgG, (F) IgM, and (G) neutralising titres. (H) S1-reactive antibodies measured at a single dilution in sera collected prior to recruitment from 47 patients with confirmed SARS-CoV-2 infection (n=10), suspected SARS-CoV-2 infection (n=16) and recruited during routine care (n=21). Antibody titres were measured in serial samples if available (range: 1-6). Dots denote patients with subsequent SARS-CoV-2 infection, triangles patients without subsequent SARS-CoV-2 infection.

**Figure S3: Gating strategies (related to Figure 3**,**4**,**5)**

(A) Gating strategy for flow analysis of S-reactive IgG, IgG, IgA. (B) Gating strategy for flow analysis of T-cell activation.

**Figure S4: Correlation of humoral and T-cell response with each other and clinical characteristics in cancer patients (related to Figure 3**,**4**,**5)**

(A) Correlation matrix of immune response in cancer patients with demographic, COVID-19 severity, and cancer characteristics. Correlation of S1-reactive antibody titres with (B) WHO severity score and (C) age. (D) Sars-CoV-2-specific T-cells in patients that received checkpoint inhibition (CPI) therapy within the three months prior to SARS-CoV-2 infection. (E) Correlation matrix of immune response in HCWs with demographics, and presence or number of symptoms. P-values < 0.05 were considered significant, significant correlations are indicated by asterisks in the correlation matrix. P-values were corrected by Benjamini-Hochberg adjustment for multiple testing. Gray squares indicate borderline significance.

**Figure S5: Antibody response in HCWs (related to Figure 4)**

S-reactive (A) IgG, (B) IGM, AND (C) IgA cells in HCWs with confirmed SARS-CoV-2 infection and/or SARS-CoV-2 antibodies (n=22), no indication of SARS-CoV-2 infection (n=51). Correlation of S1-reactive antibody titres with (B) IgG, (D) IgM, and (E) neutralising titres.

**Figure S6: T-cell response in cancer patients and HCWs (related to Figure 5)**

Comparison of CD4^+^ and CD8^+^ T-cells numbers in (A) cancer patients vs (B) HCWs. Significance was tested by Wilcoxon signed rank test, ** p < 0.01, *** p < 0.001. (C) Correlation matrix of IFN-11 secretion, Sars-CoV-2-specific CD4^+^ and CD8^+^ T-cells. Significance is indicated by asterisks, p-values were adjusted for multiple testing using Benjamini Hochberg correction. Correlation of S1-reactive antibody titres with Sars-CoV-2-specific (D) CD4^+^ and (E) CD8^+^ T-cells in cancer patients versus HCWs. CD4^+^ T-cells after stimulation with (F) S peptide pool, (G) M peptide pool, and (H) N peptide pool. Per patient increase in (I) CD4^+^ and (J) CD8^+^ SsT-cells after stimulation in COVID-19 patients and healthy controls

**Table S1: Baseline characteristics of CAPTURE participants, including cancer patients with suspected COVID-19 (Group A), cancer patients recruited in routine care (Group B) and healthcare worker volunteers.**

**Table S2: Baseline characteristics and clinical course in laboratory confirmed SARS-CoV-2 patients, patients with clinical suspicion of COVID-19 but negative for SARS-CoV-2, and patients recruited in routine clinical care**.

**Table S3: Baseline characteristics and work environment of healthcare worker volunteers**

**Table S4: Patient and healthcare volunteer shielding practices and potential sources of transmission**

**Table S5: Clinical summary of ICU stay in 4 patients with confirmed SARS-CoV2 infection**

**Table S6: Univariate logistic regression analysis of severe Covid-19 outcomes and neutralising response in cancer patients**

**Table S7: Univariate logistic regression analysis of symptomatic COVID-19 and neutralising response in HCWs**

**Table S8: Comparison of clinical characteristics between confirmed positive cancer patients and HCWs**

## Notes

### Clinical Trial

NCT03226886

### Author Declarations

CAPTURE was approved as a substudy of TRACERx Renal (NCT03226886). TRACERx Renal was initially approved by NRES Committee London Fulham on 17/01/2012. The TRACERx Renal sub-study CAPTURE was submitted as part of Substantial Amendment 9 and approved by London - Fulham Research Ethics Committee on 01/05/2020 and the Health Research Authority on 30/04/2020. CAPTURE is performed in accordance with the ethical principles in the Declaration of Helsinki, Good Clinical Practice and applicable regulatory requirements.

