## Supplemental Material for "Adaptive immunity to SARS-CoV-2 in cancer patients: The CAPTURE study"

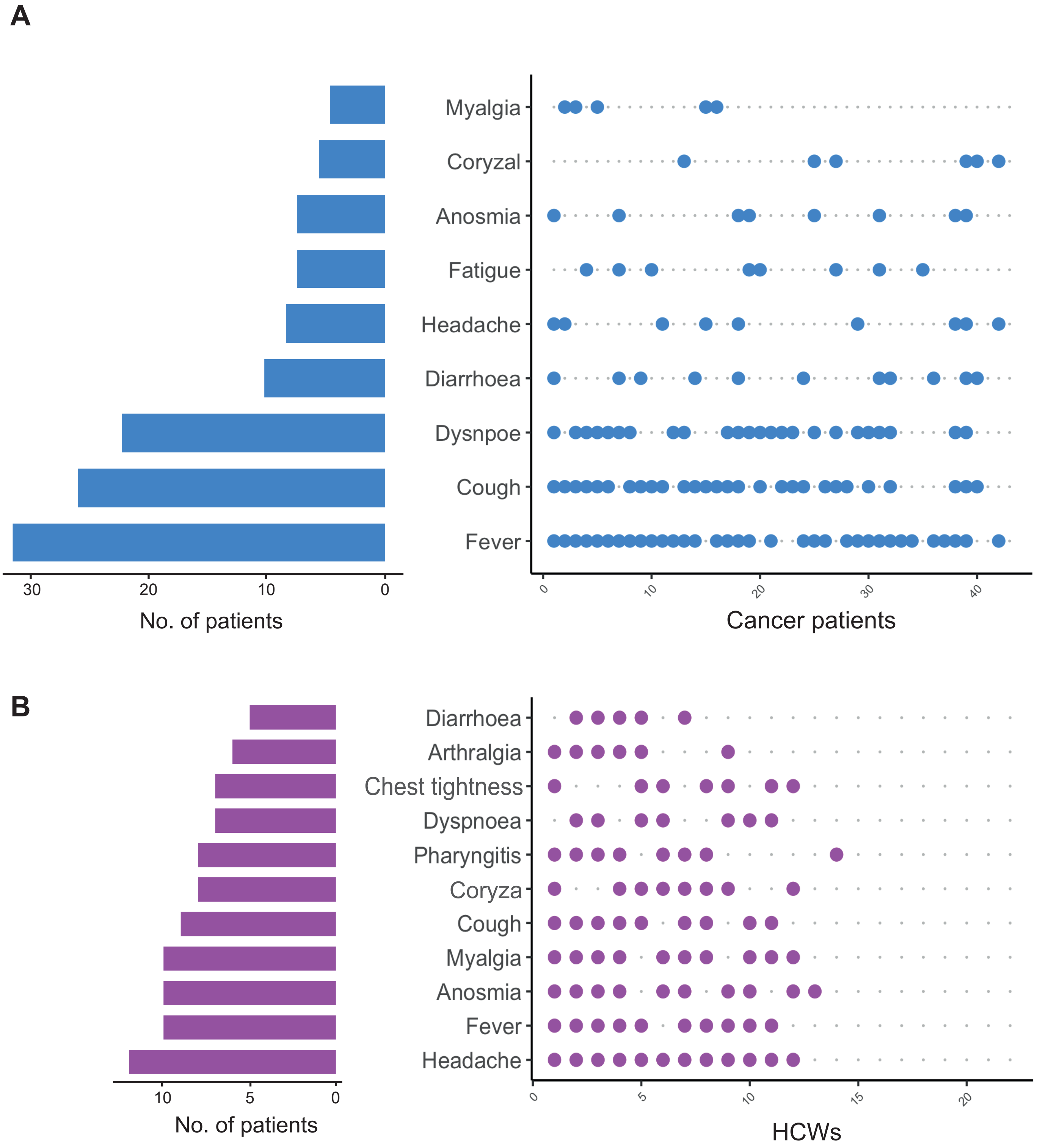
A
Myalgia
Coryzal
Anosmia
Fatigue
Headache
Diarrhoea
Dysnpoe
Cough
Fever
30
20
10
0
No. of patients
0
10
20
30
40
Cancer patients
B
Diarrhoea
Arthralgia
Chest tightness
Dyspnoea
Pharyngitis
Coryza
Cough
Myalgia
Anosmia
Fever
Headache
10
5
0
No. of patients
0
5
10
15
20
HCWs

Figure S1

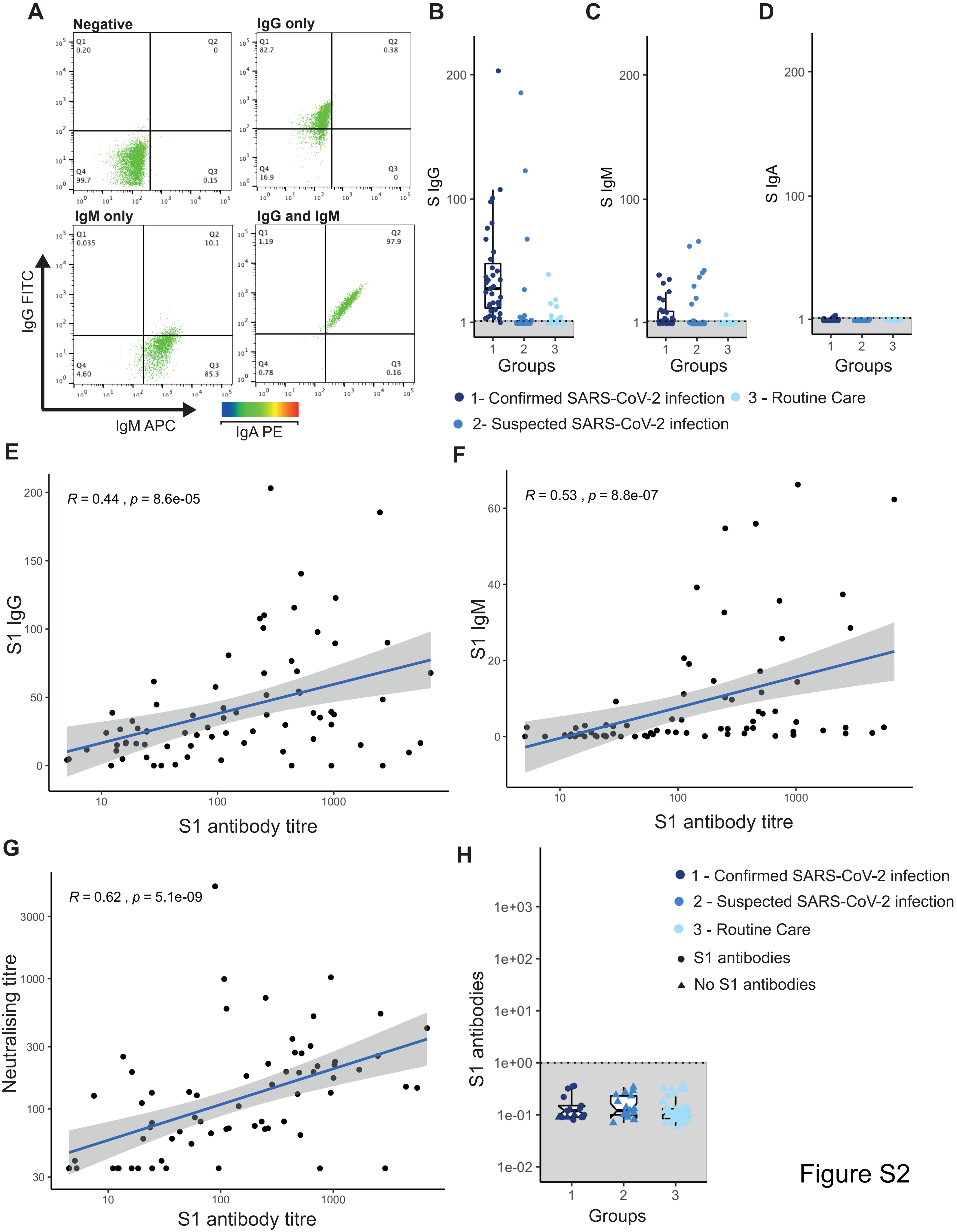
A
Negative
IgG only
IgM only
IgG and IgM
Q1 0.20
Q2 0
Q3 0.15
Q4 99.7
Q1 82.7
Q2 0.38
Q3 0
Q4 16.9
Q1 0.035
Q2 10.1
Q3 85.3
Q4 4.60
Q1 1.19
Q2 97.9
Q3 0.16
Q4 0.78
IgG FITC
IgM APC
IgA PE
B
C
D
S IgG
S IgM
S IgA
Groups
1- Confirmed SARS-CoV-2 infection
2- Suspected SARS-CoV-2 infection
3 - Routine Care
E
R = 0.44 , p = 8.6e-05
S1 IgG
S1 antibody titre
F
R = 0.53 , p = 8.8e-07
S1 IgM
S1 antibody titre
G
R = 0.62 , p = 5.1e-09
Neutralising titre
S1 antibody titre
H
S1 antibodies
Groups
1 - Confirmed SARS-CoV-2 infection
2 - Suspected SARS-CoV-2 infection
3 - Routine Care
S1 antibodies
No S1 antibodies

Figure S2

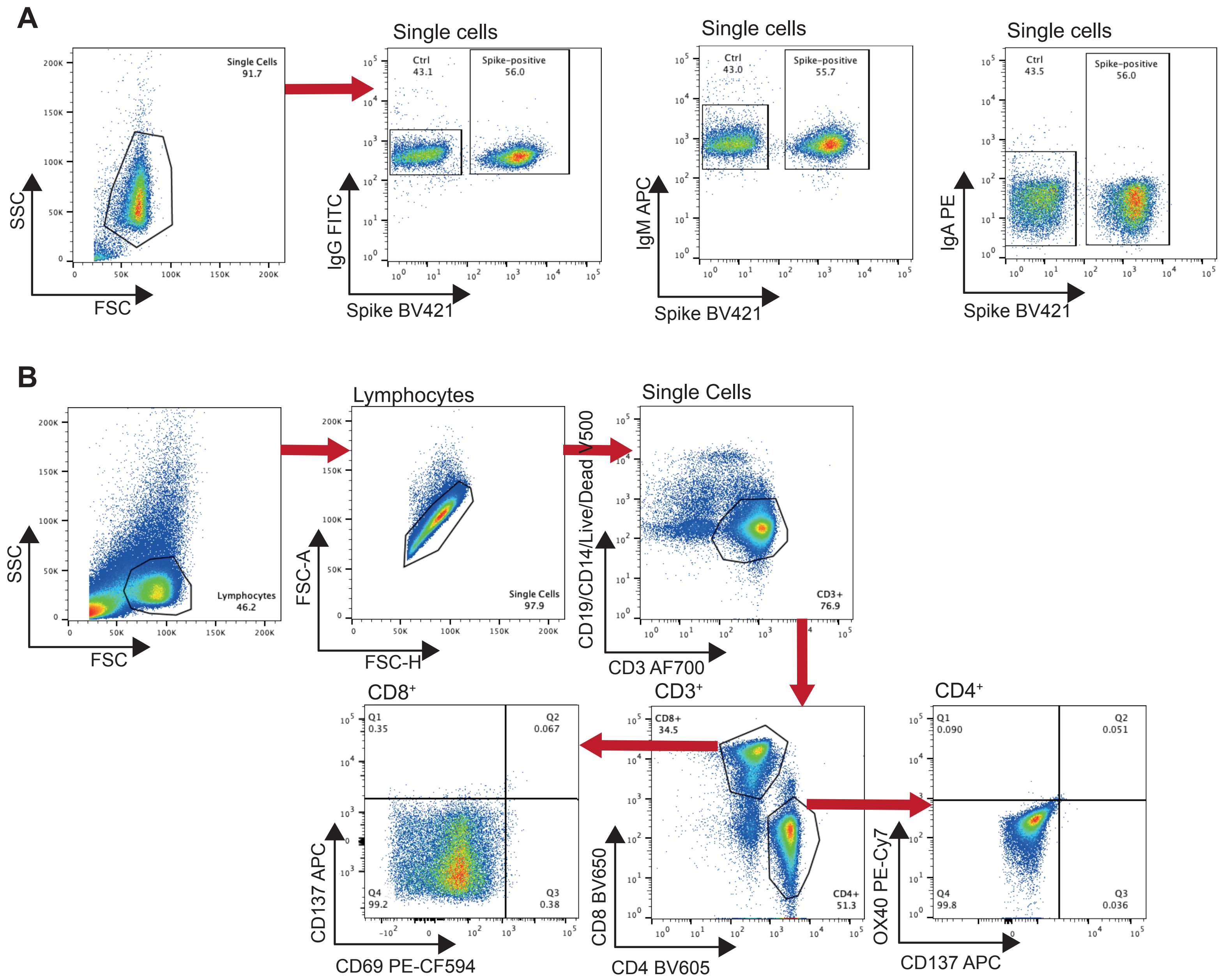
A
Single Cells 91.7
SSC
FSC
Single cells
Ctrl 43.1
Spike-positive 56.0
IgG FITC
Spike BV421
Single cells
Ctrl 43.0
Spike-positive 55.7
IgM APC
Spike BV421
Single cells
Ctrl 43.5
Spike-positive 56.0
IgA PE
Spike BV421
B
Lymphocytes 46.2
SSC
FSC
Lymphocytes
Single Cells 97.9
FSC-A
FSC-H
Single Cells
CD3+ 76.9
CD19/CD14/Live/Dead V500
CD3 AF700
CD8⁺
Q1 0.35
Q2 0.067
Q4 99.2
Q3 0.38
CD137 APC
CD69 PE-CF594
CD3⁺
CD8+ 34.5
CD4+ 51.3
CD8 BV650
CD4 BV605
CD4⁺
Q1 0.090
Q2 0.051
Q4 99.8
Q3 0.036
OX40 PE-Cy7
CD137 APC

Figure S3

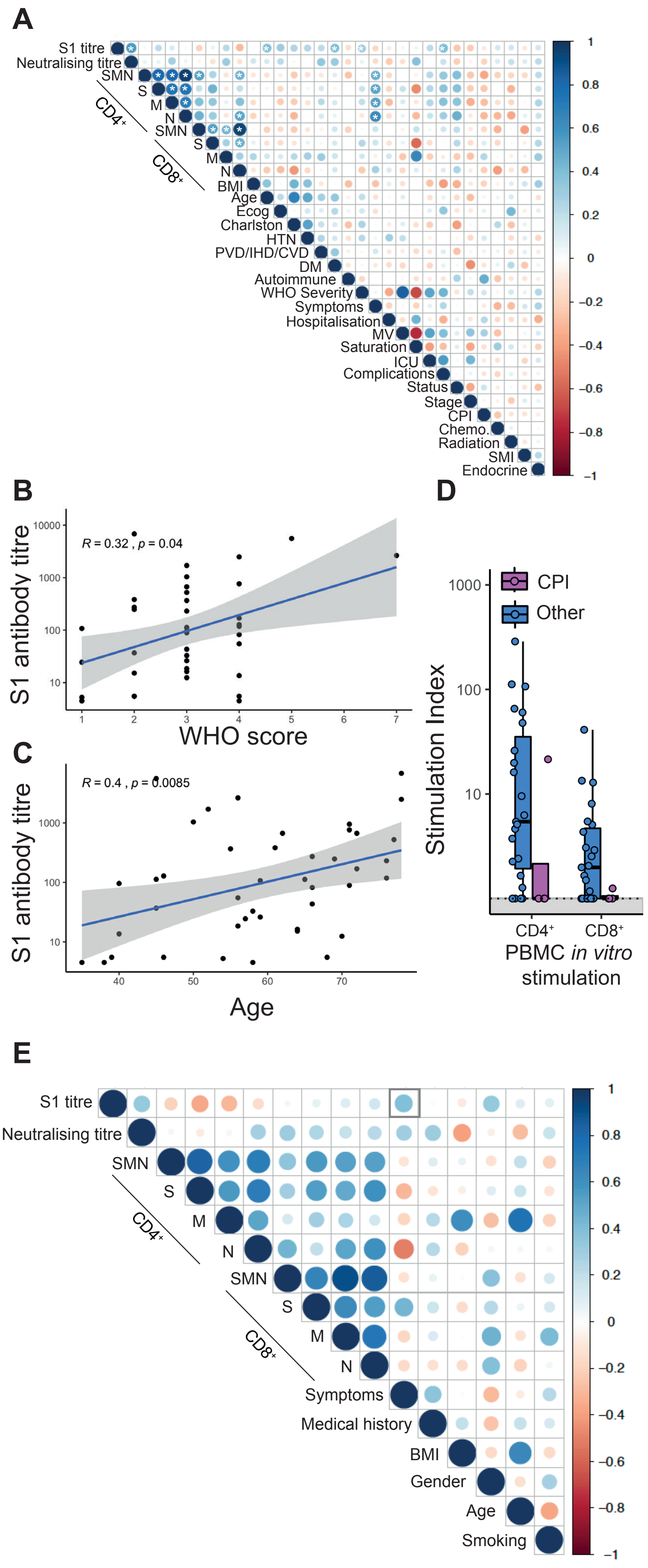

A
S1 titre
Neutralising titre
SMN
S
M
N
CD4+
SMN
S
M
N
CD8+
BMI
Age
Ecog
Charlston
HTN
PVD/IHD/CVD
DM
Autoimmune
WHO Severity
Symptoms
Hospitalisation
MV
Saturation
ICU
Complications
Status
Stage
CPI
Chemo.
Radiation
SMI
Endocrine
B
R = 0.32 , p = 0.04
S1 antibody titre
WHO score
C
R = 0.4 , p = 0.0085
S1 antibody titre
Age
D
CPI
Other
Stimulation Index
CD4+
CD8+
PBMC in vitro stimulation
E
S1 titre
Neutralising titre
SMN
S
M
N
CD4+
SMN
S
M
N
CD8+
Symptoms
Medical history
BMI
Gender
Age
Smoking


Figure S4

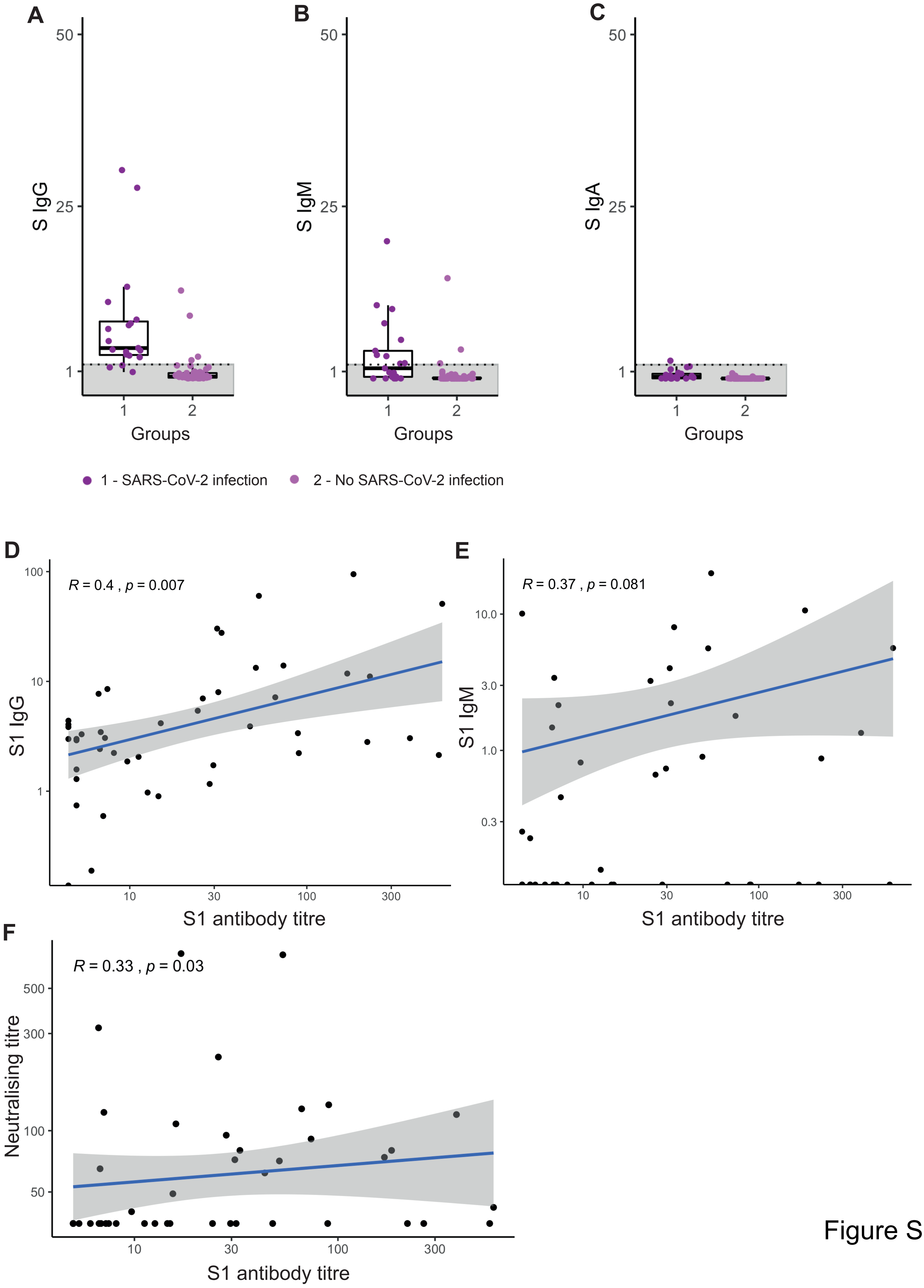
A
B
C
S IgG
S IgM
S IgA
50
25
1
Groups
1 - SARS-CoV-2 infection
2 - No SARS-CoV-2 infection
D
E
F
R = 0.4 , p = 0.007
R = 0.37 , p = 0.081
R = 0.33 , p = 0.03
S1 IgG
S1 IgM
Neutralising titre
S1 antibody titre

Figure S5

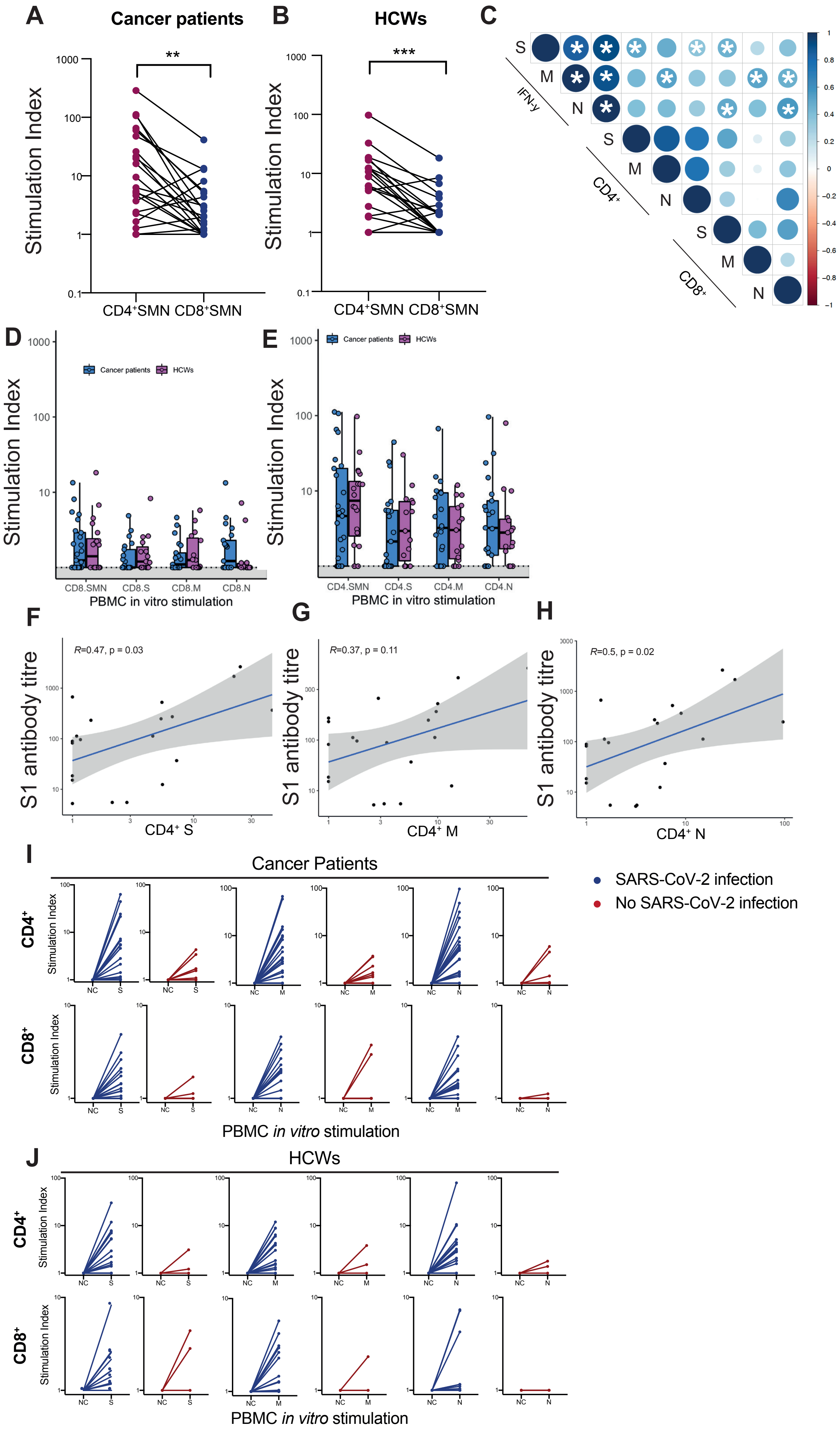
A
Cancer patients
Stimulation Index
CD4+SMN CD8+SMN
B
HCWs
C
IFN-γ
CD4+
CD8+
D
Cancer patients
HCWs
PBMC in vitro stimulation
E
F
R=0.47, p = 0.03
S1 antibody titre
CD4+ S
G
R=0.37, p = 0.11
CD4+ M
H
R=0.5, p = 0.02
CD4+ N
I
Cancer Patients
SARS-CoV-2 infection
No SARS-CoV-2 infection
J
HCWs

Figure S6

**Table S1**: Baseline characteristics of CAPTURE participants, including cancer patients with suspected COVID-19 (Group A), cancer patients recruited in routine care (Group B) and healthcare worker volunteers.

| | **Groups A, B** | **Group A** | **Group B** | **Group C** | |
|---|---|---|---|---|---|
| **Cohort Characteristics, n (%)[a]** | n =144[a] | n=74 | n=70 | n = 64/73 | p-value[b] |
| Age | | | | | |
| 18-35 | 7 (5) | 4 (5) | 3 (4) | 25 (39) | |
| 36-45 | 15 (10) | 10 (14) | 5 (7) | 18 (24) | **<0.001** |
| 46- 65 | 27 (19) | 12 (16) | 15 (21) | 17 (27) | |
| >65 | 95 (66) | 48 (65) | 47 (67) | 4 (6) | |
| Male | 69 (48) | 34 (46) | 35 (50) | 15 (23) | **<0.01** |
| **Cancer diagnosis** | | | | | |
| Skin | 38 (26) | 2 (3) | 36 (51) | - | |
| Urology | 27 (19) | 6 (8) | 21 (30) | - | |
| Gastrointestinal | 23 (16) | 22 (30) | 1 (1) | - | |
| Breast | 15 (10) | 15 (20) | 0 (0) | - | **<0.001** |
| Haematological | 10 (7) | 9 (12) | 1 (1) | - | |
| Gynaecological | 9 (6) | 4 (5) | 5 (7) | - | |
| Lung | 9 (6) | 8 (11) | 1 (1) | - | |
| Sarcoma | 9 (6) | 4 (5) | 5 (7) | - | |
| Other[c] | 4 (3) | 4 (5) | 0 (0) | - | |
| **Cancer stage[d]** | | | | | |
| Stage I-II | 5 (4) | 4 (5) | 1 (1) | - | |
| Stage III | 30 (21) | 10 (14) | 20 (29) | - | 0.06 |
| Stage IV | 99 (69) | 51 (69) | 48 (69) | - | |
| Haematological | 10 (7) | 9 (12) | 1 (1) | - | |
| **Duration of Follow up, days, median (IQR)** | 135 (104- 159) | 148.5 (112- 177) | 127.5 (104- 202) | 82(55-97) | **<0.001** |

a, Expressed as number and percent of affected individuals unless otherwise specified. Denominators reflect available data; unless specified, unknowns are not included; b, Significance tests: Chi-squared test of categorical variables and Mann-Whitney U test for comparison of median values. Significance threshold p<0.05.; c, Other cancer types include: head and neck cancers (n=2), glioblastoma, thymic carcinoma; d, Cancer stage at enrolment according to AJCC stage definition. IQR, interquartile range.

**Table S2**: Baseline characteristics and clinical course in laboratory confirmed SARS-CoV-2 patients, patients with clinical suspicion of COVID-19 but negative for SARS-CoV-2, and patients recruited in routine clinical care.

| | Cancer patients | Group A + B SARS-Cov2 positive[a] | Group A SARS-CoV2[a] negative | Group B SARS-CoV negative[a] | |
|---|---|---|---|---|---|
| | n= 144[b] | n= 43 | n= 33 | n= 68 | p-value[c] |
| **DEMOGRAPHICS** | | | | | |
| Age, years (median, IQR) | 58.9 (28-82) | 59.4 (38- 78) | 57.6 (28- 78) | 59.1 (28- 82) | 0.954 |
| Male, n (%) | 69 (47.9) | 21 (48.9) | 14 (42.4) | 34 (50) | 0.766 |
| **PAST MEDICAL HISTORY** | | | | | |
| HTN | 38 (26) | 10 (24)) | 5 (15) | 23 (33) | 0.182 |
| PVD/IHD/CVD | 12 (8) | 3 (7) | 6 (18) | 3 (4) | 0.058 |
| Diabetes Mellitus | 15 (10) | 8 (20) | 2 (6) | 5 (7) | 0.057 |
| Obesity (BMI ≥30) | 24 (17) | 11 (26) | 8 (24) | 25 (17) | 0.308 |
| Inflammatory/autoimmune | 11 (8) | 1 (2) | 3 (9) | 7 (10) | 0.677 |
| **Smoking History** | | | | | |
| Current smoker | 44 (31) | 15 (37) | 12 (36) | 17 (24) | |
| Ex-smoker | 71 (49) | 18 (44) | 12 (36) | 41 (59) | 0.474 |
| Never smoker | 22 (15) | 5 (12) | 7 (21) | 10 (14) | |
| Unknown | 7 (5) | 3 (7) | 2 (6) | 2 (3) | |
| **ONCOLOGICAL HISTORY** | | | | | |
| **Cancer diagnosis, n (%)** | | | | | |
| Skin | 38 (26) | 0 (0) | 2 (6) | 36 (51) | |
| Urology | 27 (19) | 4 (10) | 2 (6) | 21 (30) | |
| Gastrointestinal | 23 (16) | 13 (32) | 9 (27) | 1 (1) | |
| Breast | 15 (10) | 8 20) | 7 (21) | 0 (0) | **<0.001** |
| Haematological | 10 (7.0) | 5 (12) | 4 (12) | 1 (1) | |
| Gynaecological | 9 (6) | 2 (5) | 2 (6) | 5 (7) | |
| Lung | 9 (6) | 5 (12) | 3 (9) | 1 (1) | |
| Sarcoma | 9 (6) | 1 (2) | 3 (9) | 5 (7) | |
| Other | 4 (3) | 3 (7) | 1 (3) | 0 (0) | |

| | | | | | |
|---|---|---|---|---|---|
| **Cancer stage[d], n (%)** | | | | | |
| Stage I-II | 5 (4) | 2 (5) | 2 (6) | 1 (1) | |
| Stage III | 30 (21) | 7 (17) | 3 (9) | 20 (29) | 0.056 |
| Stage IV | 99 (69) | 27 (66) | 24 (72) | 48 (69) | |
| Haematological | 10 (7) | 5 (12) | 4 (12) | 1 (1) | |
| **Treatment within 12 weeks, n (%)** | | | | | |
| SACT, any | 113 (78) | 35 (81) | 27 (82) | 51 (75) | |
| Chemotherapy | 57 (31) | 28 (46) | 22 (49) | 7 (9) | |
| Small Molecule Inhibitor | 41 (22) | 12 (20) | 5 (11) | 5 (11) | |
| Anti-PD1/PDL1 | 22 (12) | 1 (2) | 3 (7) | 18 (23) | |
| Anti-PD1 & anti-CTLA4 | 15 (8) | 1 (2) | 1 (2) | 13 (17) | **<0.001** |
| CPI combination, other | 9 (5) | 2 (3) | 1 (2) | 6 (8) | |
| Radiotherapy | 18 (10) | 7 (12) | 8 (18) | 3 (4) | |
| Surgery | 8 (4) | 2 (3) | 4 (9) | 2 (3) | |
| Endocrine Therapy | 6 (3) | 5 (8) | 1 (2) | 0 (0) | |
| None | 5 (3) | 1 (2) | 0 (0) | 4 (5) | |
| Other | 4 (2) | 2 (3) | 0 (0) | 2 (3) | |
| **Cancer status[e], n (%)** | | | | | |
| **Solid cancer, advanced (n = 99)** | **n = 99** | **n = 27** | **n = 24** | **n = 48** | |
| Resected, NED | 7 (7) | 1 (4) | 0 (0) | 6 (13) | |
| SACT, CR/PR | 29 (29) | 5 (19) | 3 (13) | 21 (44) | |
| SACT, SD | 32 (32) | 14 (52) | 6 (25) | 12 (25) | **<0.001** |
| SACT, PD | 31 (31) | 7 (26) | 15 (62.5) | 9 (19) | |
| **Solid cancer, localised (n=35)** | **n = 35** | **n = 9** | **n = 5** | **n = 21** | |
| Resected, NED | 8 (23) | 3 (33) | 1 (20) | 4 (19) | |
| (Neo-) adjuvant SACT | 17 (49) | 2 (22) | 3 (60) | 12 (57) | |
| Radical CRT | 1 (3) | 1 (11) | 0 (0) | 0 (0) | 0.731 |
| SACT, CR/PR | 5 (14) | 1 (11) | 1 (20) | 3 (14) | |
| SACT, SD | 2 (6) | 1 (11) | 0 (0) | 1 (5) | |
| SACT, PD | 2 (6) | 1 (11) | 0 (0) | 1 (5) | |
| **Haematological, (n= 10)** | **n = 10** | **n = 5** | **n = 4** | **n = 1** | |
| SACT, in remission | 5 (50) | 3 (60) | 1 (25) | 1 (100) | |
| SACT, stable active disease | 2 (20) | 0 (0) | 2 (50) | 0 (0) | 0.331 |

| | | | | | |
|---|---|---|---|---|---|
| SACT, progressive disease | 3 (30) | 2 (40) | 1 (25) | 0 (0) | |
| **CLINICAL OUTCOMES & IMPACT** | | | | | |
| Duration of follow-up, median (IQR) | 135 (104- 159) | 147 (96- 153) | 177 (126- 182) | 104 (68- 149) | **0.001** |
| **At data cutoff** | | | | | |
| Deceased, yes | 17 (12) | 4 (10) | 12 (36) | 1 (1) | **<0.001** |
| Primary cause death: | | | | | |
| Progressive Cancer | 15 (10) | 4 (10) | 10 (30) | 1 (1) | |
| Complications of COVID-19[f] | 0 (0) | 0 (0) | 0 (0) | 0 (0) | 0.623 |
| Other | 2 (1) | 0 (0) | 2 (6) | 0 (0) | |
| **Impact on cancer treatment** | | | | | |
| Delay of treatment | 44 (31) | 26 (61) | 5 (15) | 14 (20) | |
| Dose modification or change of therapy | 8 (6) | 4 (9) | 2 (6) | 2 (3) | **<0.001** |
| Premature discontinuation | 1 (1) | 0 (0) | 0 (0) | 1 (1) | |

a, Indicates positive/negative for SARS-CoV2 by laboratory definition; b, Expressed as number and percent of affected individuals unless otherwise specified. Denominators reflect available data; unless specified, unknowns are not included; c, Significance tests: Chi-squared test of categorical variables and Mann-Whitney U test for comparison of median values; d, Cancer stage at enrolment according to AJCC stage definition; e, Cancer status as defined by RECIST v2.2 criteria; f, One patient dies within 30 days of PCR positivity, but not due to COVID-19

BMI, body mass index; CPI, checkpoint inhibitor; CR, complete response; CRT, chemoradiotherapy; CTLA-4, cytotoxic T-lymphocyte-associated protein 4; HTN, hypertension; PD, progressive disease; PR, partial response; PVD, peripheral vascular disease; IHD, ischaemic heart disease; CVD, cerebrovascular disease; IQR, interquartile range; NED, no evidence of disease; SACT, systemic anti-cancer therapy; SD, stable disease; PD1, programmed death 1; PDL1, programmed death ligand 1.

**Table S3:** Baseline characteristics professional role and work environment of healthcare volunteers

| | Group C, n=73 | | | |
|---|---|---|---|---|
| | All | SARS-CoV2 +[a] | SARS-CoV2-[a] | |
| **PROFILES Completion** | n = 64/73[b] | n= 20/22 | n= 44/51 | p-value[c] |
| Age | | | | |
| 18-35 | 25 (39) | 5 (20) | 20 (45.5) | |
| 36-45 | 18 (24) | 7 (80) | 11 (25.0) | 0.06 |
| 46- 65 | 17 (27) | 5 (25) | 12 (45.4) | |
| >65 | 4 (6) | 3 (15) | 1 (2.3) | |
| Male | 15 (23) | 4 (20) | 9 (20.5) | 1 |
| Smoking | | | | |
| Current | 1 (2) | 0 (0) | 1 (2.3) | |
| Previous | 18 (28) | 6 (30) | 12 (27.2) | 0.782 |
| Never | 35 (55) | 14 (70) | 31 (70.4) | |
| **PAST MEDICAL HISTORY** | n= 63 | n=19 | n = 44 | |
| prev_illness, yes | 21 (33) | 5 (26.3) | 16 (36.4) | 0.627 |
| Obese | 11 (17) | 6 (31.6) | 5 (11.6) | 0.114 |
| HTN | 2 (3) | 0 (0) | 2 (4.5) | 0.871 |
| CAD | 0 (0) | 0 (0) | 0 (0) | - |
| Stent | 0 (0) | 0 (0) | 0 (0) | - |
| Hypercholesterolaemia | 2 (3) | 0 (0) | 2 (4.5) | 0.871 |
| CVD | 0 (0) | 0 (0) | 0 (0) | - |
| DVT | 0 (0) | 0 (0) | 0 (0) | - |
| PE | 0 (0) | 0 (0) | 0 (0) | - |
| NIDMM | 0 (0) | 0 (0) | 0 (0) | - |
| IDMM | 0 (0) | 0 (0) | 0 (0) | - |
| Hyperthyroidism | 0 (0) | 0 (0) | 0 (0) | - |
| Hypothyroidism | 4 (6) | 1 (5.2) | 3 (6.8) | 1 |
| CKD | 0 (0) | 0 (0) | 0 (0) | - |
| COPD | 0 (0) | 0 (0) | 0 (0) | - |
| Asthma | 4 (6) | 1 (5.2) | 3 (6.8) | 1 |
| **ROLE & ENVIRONMENT** | n = 64 | n = 20 | n = 44 | |
| Patient facing role, yes | 58 (91) | 18 (90) | 40 (91) | 1 |
| Work environment | | | | |
| ICU | 8 (13) | 6 (30) | 2 (5) | **0.014** |
| Theatres | 2 (3) | 0 (0) | 2 (5) | 0.846 |
| Inpatients/ward | 30 (47) | 9 (45) | 21 (48) | 1 |
| Outpatients | 37 (58) | 11 (55) | 26 (59) | 1 |
| Other | 4 (6) | 2 (10) | 2 (5) | 0.972 |
| COVID cohort area, yes | 34 (53) | 8 (44.4) | 26 (65) | 0.694 |
| Aerosol procedure, yes | 8 (13) | 3 (16.7) | 5 (13) | 0.989 |
| Close colleague, positive | 39 (61) | 11 (55) | 28 (64) | 0.703 |
| Off-work isolation, yes | 29 (45) | 14 (70) | 15 (34) | **0.016** |
| **SYMPTOMS** | n = 64 | n=20 | n=44 | |
| Fever | 16 (25) | 10 (50) | 8 (18) | **0.035** |
| Headache | 23 (36) | 12 (60) | 14 (32) | 1 |
| Coryzal illness | 16 (25) | 8 (40) | 11 (25) | 0.860 |
| Pharyngitis | 19 (30) | 7 (35) | 12 (27) | 1 |
| Anosmia | 12 (19) | 10 (50) | 3 (7) | **<0.001** |
| Cough | 18 (28) | 9 (45) | 10 (23) | 0.356 |

| | | | | |
|---|---|---|---|---|
| Dyspnoea | 7 (11) | 7 (35) | 3 (7) | **0.028** |
| Chest tightness | 7 (11) | 7 (35) | 2 (5) | 0.057 |
| Diarrhoea | 7 (11) | 5 (25) | 3 (7) | 0.769 |
| Myalgia | 15 (23) | 10 (50) | 6 (14) | **0.035** |
| Arthralgia | 7 (11) | 6 (30) | 2 (5) | **0.037** |
| Rash | 1 (2) | 0 (0) | 1 (2) | 1 |

a, Indicates positive/negative for SARS-CoV2 by laboratory definition ; b, 64 out of 73 participants in the healthcare worker completed baseline PROFILES questionnaire. Expressed as number and percent of affected individuals unless otherwise specified. Denominators reflect available data; unless specified, unknowns are not included; c, Significance tests: Chi-squared test of categorical variables and Mann-Whitney U test for comparison of median values. Significance threshold $p<0.05$; d, Volunteers may work in >1 environment. CAD, coronary artery disease; CKD, chronic kidney disease; COPD, chronic obstructive pulmonary disease; CVD, cerebrovascular disease; HTN, hypertension; ICU, intensive care unit

**Table S4: Patient and healthcare volunteer shielding practices and potential sources of transmission**

| | Group A & B, n=144 | | | Group C, n= 73 | | |
|---|---|---|---|---|---|---|
| | SARS-CoV2 +[a] | SARS-CoV2 -[a] | | SARS-CoV2 +[a] | SARS-CoV2 -[a] | |
| **PROFILES Completion** | | | p-value | | | p-value[b] |
| **LIFESTYLE & SHIELDING** | n = 20[c] | n = 74[c] | | n=19[c] | n=44[c] | |
| Advised to shield, yes | 19 (85) | 63 (85) | 0.426 | - | - | - |
| Time shielding | | | | | | |
| 0 days | 3 (15) | 15 (2) | | - | - | - |
| 1-14 days | 6 (30) | 2 (3) | | - | - | - |
| 14-28 days | 0 (0) | 2 (3) | **0.002** | - | - | - |
| >28 days | 11 (55) | 55 (74) | | - | - | - |
| Household size[d] | | | | | | |
| 1 | 1 (5) | 18 (24) | | 5 (26) | 9 (21) | |
| 2-3 | 10 (50) | 38 (51) | 0.075 | 9 (47) | 25 (57) | 0.886 |
| >=4 | 9 (45) | 18 (24) | | 5 (26) | 10 (24) | |
| Household positive contact, yes | 6 (30) | 0 (0) | **<0.001** | 2 (11) | 1 (2) | 0.442 |

a, Indicates positive/negative for SARS-CoV2 by laboratory definition; b, Significance tests: Chi-squared test of categorical variables and Mann-Whitney U test for comparison of median values. Significance threshold p<0.05; c, Expressed as number and percent of affected individuals unless otherwise specified. Denominators reflect available data; unless specified, unknowns are not included; d, Household size inclusive of participant.

**Table S5: Clinical summary of ICU stay in 4 patients with confirmed SARS-CoV2 infection**

| Patient | Cancer Diagnosis | Time in ICU (days) | Respiratory support | Reason for ICU admission | Requirement for inotropes |
|---|---|---|---|---|---|
| CV0008 | T-ALL | 1 | Oxygen (nasal prongs, mask) | In context of COVID-19: hypoxia, 8 days after having tested positive for SARS-CoV-2, also received tocilizumab at that time | No |
| CV0004 | Spindle cell sarcoma | 1 | Oxygen (nasal prongs, mask) | Not in context of COVID-19: post-op post VATS | No |
| CV0037 | DLBCL | 3 | Oxygen (nasal prongs, mask) | In context of COVID: hypoxia, 1 day after having tested positive for SARS-CoV-2, also received tocilizumab at that time | No |
| CV0025 | Breast cancer | 100 | Oxygen, NIV, mechanical ventilation (2 episodes: 1st 59 days, 2nd 30 days) | In context of COVID: hypoxia, tachypnoeic, 6 days after having tested positive for SARS-CoV-2, received noradrenaline | Yes |

DLBCL, diffuse large B-cell lymphoma; ICU, intensive care unit; NIV, non-invasive ventilation; T-ALL, T-cell acute lymphoblastic leukaemia; VATS, video assisted thoracoscopic surgery

**Table S6: Univariate logistic regression analysis of severe COVID-19 outcomes and neutralizing response in cancer patients**

| Parameter | Severe COVID-19 | | Hospital admission | | ICU admission | | Complications | | Neutralizing Antibodies | |
|---|---|---|---|---|---|---|---|---|---|---|
| | **OR (CI)** | **p-value** | **OR (CI)** | **p-value** | **OR (CI)** | **p-value** | **OR (CI)** | **p-value** | **OR (CI)** | **p-value** |
| Age | 1.01(0.96-1.08) | 0.59 | 1.02(0.97-1.07) | 0.54 | 0.95(0.85-1.05) | 0.35 | 0.90(0.16-4.29) | 054 | 1.04 (0.97-1.11) | 0.28 |
| BMI | 1.02(0.91-1.14) | 0.73 | 0.99(0.89-1.10) | 0.91 | 0.85(0.57-1.08) | 0.29 | **0.70(0.48-0.89)** | **0.02** | 1.16(0.99-1.44) | 0.11 |
| ECOG | 1.39(0.47-4.33) | 0.55 | 2.15(0.77-6.76) | 0.16 | 1.61(0.24-11.62) | 0.62 | **4.42(1.16-23.42)** | **0.046** | 0.84(0.24-1.94) | 0.79 |
| Gender | 1.49(0.39-6.04) | 0.56 | 1.92(0.57-6.72) | 0.29 | NA | NA | 1.76(0.37-9.69) | 0.48 | 1.00(0.21-4.87) | 1.00 |
| Smoking | 1.11(0.50-2.49) | 0.80 | 1.35(0.66-2.86) | 0.42 | 2.92(0.66-21.17) | 0.21 | 1.10(0.44-2.79) | 0.85 | 0.94(0.37-2.35) | 0.89 |
| Cancer diagnosis | 1.22(0.96-1.60) | 0.17 | 1.06(0.85-1.32) | 0.68 | 1.33(0.86-2.43) | 0.31 | 0.98(0.67-1.37) | 0.92 | 1.01(0.77-1.35) | 1.00 |
| Solid vs. hematological malignancy | 2.9(0.31-26.93) | 0.32 | 2.57(0.30-54.54) | 0.43 | **3.8(2.65-1090)** | **0.01** | 5.50(0.57-53.98) | 0.12 | 0.18(0.02-1.80) | 0.12 |
| Cancer status | 1.00(0.57-1.67) | 0.99 | 0.74(0.43-1.20) | 0.24 | 0.94(0.29-2.15) | 0.90 | 1.26(0.69-2.24) | 0.42 | 0.95(0.54-1.81) | 0.86 |
| Stage | 3.39(1.03-16.17) | 0.07 | 2.12(0.83-6.51) | 0.14 | 3.00(0.19-376.20) | 0.65 | 1.04(0.69-2.24) | 0.95 | 0.24(0.04-0.98) | 0.08 |
| CPI, all combinations | 0.85(0.04-7.48) | 0.89 | 0.23(0.01-2.00) | 0.22 | NA | NA | NA | NA | 0.18(0.02-1.80) | 0.12 |
| Chemotherapy | 1.01(0.26-4.09) | 0.99 | 2.22(0.65-7.92) | 0.21 | NA | NA | 0.67(0.14-3.24) | 0.61 | 0.38(0.05-1.90) | 0.27 |
| Small molecule inhibitor | 0.68(0.09-3.46) | 0.67 | 0.98(0.22-4.60) | 0.99 | NA | NA | 0.48(0.02-3.33) | 0.52 | NA | NA |
| Endocrine therapy | 3.11(0.50-19.63) | 0.21 | NA | NA | 3.50(0.15-43.99) | 0.34 | 0.85(0.04-6.55) | 0.89 | 1.21(0.16-25.05) | 0.87 |
| Radiotherapy | 4.67(0.86-28.20) | 0.07 | 2.24(0.42-17.07) | 0.37 | **14.00(1.14-336.9)** | **0.047** | 4.65(0.74-28.40) | 0.09 | 0.52(0.09-4.20) | 0.49 |
| Surgery | NA | NA | 0.37(0.02-4.16) | 0.43 | NA | NA | NA | NA | NA | 0.99 |
| No. of comorbidities | 0.98(0.57-1.63) | 0.94 | 0.99(0.63-1.57) | 0.97 | 0.30(0.02-1.04) | 0.16 | 1.45(0.82-2.70) | 0.21 | 1.44(0.79-2.94) | 0.26 |
| HTN | 1.71(0.37-7.38) | 0.47 | 1.54(0.39-6.88) | 0.55 | NA | NA | 0.96(0.13-5.12) | 0.97 | 2.92(0.43-58.23) | 0.34 |
| PVD/IHD/CVD | 1.32(0.06-15.16) | 0.83 | NA | 0.99 | NA | NA | 2.36(0.10-28.22) | 0.51 | NA | 0.99 |
| Diabetes Mellitus | 0.83(0.11-4.37) | 0.84 | 1.40(0.30-7.71) | 0.67 | NA | NA | 1.61(0.20-9.26) | 0.61 | NA | NA |
| Inflammatory/ autoimmune | NA | NA | NA | NA | NA | NA | NA | NA | 0.21(0.01-5.79) | 0.29 |
| Charlston comorbidity index | 1.05(0.83-1.36) | 0.69 | 1.15(0.93-1.46) | 0.21 | 0.71(0.41-1.10) | 0.15 | 1.04(0.79-1.40) | 0.80 | 1.10(0.83-1.46) | 0.50 |
| $spO_2$ | 0.30(0.01-0.68) | 0.11 | 0.75(0.48-0.97) | 0.09 | 0.83(0.55-1.16) | 0.25 | 0.87(0.71-1.03) | 0.13 | 1.19(0.99-1.50) | 0.07 |
| HB at diagnosis | 0.99(0.95-1.04) | 0.83 | 0.99(0.95-1.03) | 0.62 | **0.90(0.78-0.98** | **0.049** | 0.97(0.92-1.02) | 0.21 | 0.98(0.93-1.02) | 0.37 |
| WBC at diagnosis | 0.77(0.50-1.00) | 0.14 | 0.79(0.58-1.00) | 0.12 | 0.47(0.13-0.97) | 0.13 | 0.81(0.54-1.01) | 0.25 | NA | NA |
| Neutrophils at diagnosis | 0.93(0.55-1.46) | 0.76 | 0.78(0.49-1.18) | 0.25 | 0.92(0.32-1.94) | 0.84 | 0.84(0.46-1.37) | 0.51 | 1.01(0.64-1.67) | 0.95 |
| Platelets at diagnosis | 0.99(0.98-1.00) | 0.23 | 1.00(0.99-1.01) | 0.75 | 0.97(0.94-1.00) | 0.07 | 1.00(0.99-1.00) | 0.35 | 1.00(0.99(1.01) | 1.00 |
| CRP at diagnosis | 1.01(1.00-1.02) | 0.07 | 1.02(1.00-1.06) | 0.07 | 1.01(1.00-1.02) | 0.15 | 1.01(1.00-1.01) | 0.17 | 1.00(0.99-1.02) | 0.44 |
| Creatinine at diagnosis | 0.97(0.91-1.02) | 0.27 | 0.98(0.94-1.03) | 0.47 | 0.96(0.88-1.03) | 0.25 | 0.96(0.89-1.01) | 0.15 | 1.02(0.97-1.08) | 0.53 |

| | | | | | | | | | | |
|---|---|---|---|---|---|---|---|---|---|---|
| Respiratory frequency at diagnosis | 1.38(1.06-2.15) | 0.07 | 1.06(0.91-1.32) | 0.54 | 1.36(1.06-2.19) | 0.08 | 1.09(0.93-1.33) | 0.30 | 0.90(0.72-1.06) | 0.23 |
| Heart rate at diagnosis | 0.92(0.81-1.01) | 0.14 | 0.99(0.92-1.08) | 0.83 | 0.98(0.85-1.10) | 0.79 | 0.92(0.81-1.02) | 0.15 | 1.02(0.94-1.12) | 0.67 |
| Systolic pressure at diagnosis | 0.93(0.86-0.99) | 0.07 | 1.00(0.96-1.05) | 0.85 | 0.90(0.74-1.01) | 0.14 | 0.98(0.98-1.03) | 0.39 | 1.06(1.00-1.44) | 0.08 |
| Diastolic pressure at diagnosis | 0.91(0.79-1.00) | 0.10 | 0.98(0.91-1.05) | 0.62 | NA | NA | 0.97(0.88-1.05) | 0.43 | 0.97(0.89-1.05) | 0.46 |
| Temperature at diagnosis | 1.27(0.53-3.14) | 0.58 | 2.17(0.90-6.45) | 0.12 | 2.55(0.77-11.13) | 0.15 | 1.08(0.43-2.68) | 0.87 | 1.07(0.44-2.73) | 0.88 |
| Lactate at diagnosis | 1.58(0.45-7.73) | 0.49 | 0.5(0.85-1.05) | 0.56 | 0.12(0.00-2.53) | 0.39 | 3.06(0.77-26.71) | 0.19 | 3.36(0.63-71.44) | 0.29 |

Data are odds ratio with 97.5% CI in parentheses. ICU, Intensive Care Unit; ECOG, Eastern Cooperative Oncology Group; BMI, Body mass index; CPI, immune checkpoint inhibition: HTN, hypertension; PVD, Peripheral vascular disease ; IHD, Ischemic heart disease ; CVD, Cardiovascular disease ;spO2, ;HB, Haemoglobin; CRP, C-reactive protein, NA, No available; NS, non-significant.

**Table S7: Univariate logistic regression analysis of symptomatic COVID-19 and neutralising response in HCWs**

| Parameter | Symptoms | | Neutralizing Antibody | |
|---|---|---|---|---|
| | OR (CI) | p-value | OR (CI) | p-value |
| Age | 10.52(0.14-1.55) | 0.52 | 1.42(0.51-4.67) | 0.27 |
| BMI | 0.95(0.75-1.18) | 0.60 | 1.06(0.85-1.39) | 0.61 |
| Gender | 0.57(0.02-7.30) | 0.75 | 0.66(0.02-6.93) | 0.67 |
| Smoking | 1.67(0.17-14.47) | 0.64 | 5.40(0.59-126.79) | 0.19 |
| Work environment | 0.65(0.27-1.34) | 0.27 | 1.01(0.52-1.97) | 0.97 |
| COVID-19 area | 0.67(0.07-5.53) | 0.71 | 0.17(0.02-1.23) | 0.10 |

Data are odds ratio with 97.5% CI in parentheses. BMI, Body mass index; NA, No available.

**Table S8:** Comparison of clinical characteristics between confirmed positive cancer patients and HCWs

| | **Cancer patients, SARS-CoV2 positive[a]** | **Group C, SARS-CoV2 positive[a]** | |
|---|---|---|---|
| **Clinical Characteristics** | n= 43[b] | n= 22 | p-value[c] |
| Age | | | |
| 18-35 | 0 (0) | 5 (20) | |
| 36-45 | 8 (19) | 7 (80) | **<0.001** |
| 46- 65 | 6 (14) | 5 (25) | |
| >65 | 29 (67) | 3 (15) | |
| Male | 21 (49) | 4 (20) | **0.016** |
| **Past medical history** | | | |
| HTN | 10 (24) | 0 (0) | **<0.001** |
| PVD/IHD/CVD | 3 (7) | 0 (0) | **<0.001** |
| Diabetes Mellitus | 9 (19) | 0 (0) | **0.023** |
| Obesity, BMI>30 | 18 (42) | 6 (32) | 0.289 |
| Inflammatory/Autoimmune | 1 (2) | 0 (0) | 1 |
| **COVID-19 illness** | | | |
| **Symptoms** | n = 43/43 | n = 20/22 | |
| Fever | 34 (83) | 10 (50) | **<0.001** |
| Dyspnoea | 20 (47) | 7 (35) | 0.426 |
| Cough | 28 (65) | 9 (45) | 0.172 |
| Anosmia | 8 (19) | 10 (50) | **0.016** |
| Myalgia | 13 (13) | 10 (50) | 0.164 |
| Coryzal illness | 6 (14) | 8 (40) | **0.028** |
| Headache | 9 (21) | 12 (60) | **0.004** |
| Diarrhoea | 11 (26) | 5 (25) | 1 |
| Duration of symptoms, median (range) | 19 (1- 146) | 14 (3- 120) | 0.306 |

a, Indicates positive/negative for SARS-CoV2 by laboratory definition; b, Expressed as number and percent of affected individuals unless otherwise specified. Denominators reflect available data; unless specified, unknowns are not included; c, Significance tests: Chi-squared test of categorical variables and Mann-Whitney U test for comparison of median values. Significance threshold $p<0.05$.
